# Country-Specific Estimates of Misclassification Rates of Computer-Coded Verbal Autopsy Algorithms

**DOI:** 10.1101/2025.07.02.25329250

**Authors:** Sandipan Pramanik, Emily B. Wilson, Henry D. Kalter, Victor Akelo, Agbessi Amouzou, Robert E. Black, Dianna Blau, Ivalda Macicame, Jonathan A. Muir, Kyu Han Lee, Li Liu, Cynthia G. Whitney, Scott Zeger, Abhirup Datta

**Author notes:** Disclaimer: The findings and conclusions in this report are those of the author(s) and do not necessarily represent the views of the U.S. Centers for Disease Control and Prevention.

## Abstract

**Background:** Computer-coded verbal autopsy (CCVA) algorithms are commonly used to determine individual causes of death (COD) and population-level cause-specific mortality fractions (CSMF), but frequent COD misclassification leads to biased CSMF estimates. The VA-calibration framework [1,2] reduces bias by estimating misclassification rates from limited CHAMPS data, but it overlooks country-level variation in these rates, reducing the accuracy of CSMF estimates.

**Methods:** Utilizing CHAMPS data and the framework from [3], we estimate VA misclassification rates for three widely used CCVA algorithms (EAVA, InSilicoVA, InterVA), two age groups (neonates 0-27 days and children 1-59 months), and eight countries (Bangladesh, Ethiopia, Kenya, Mali, Mozambique, Sierra Leone, South Africa, other). We then use the Mozambique-specific rates to calibrate VA-only data from the COMSA project in Mozambique.

**Findings:** We report three key findings. First, the country-specific model better fits CHAMPS misclassification rates than the homogeneous model, reducing average absolute loss by 34-38% for neonates and 13-24% for children. Second, CCVA algorithms show consistent misclassification patterns, systematically over- or underestimating certain causes. Third, calibrating COMSA data increases neonatal CSMF for sepsis/meningitis/infection and decreases it for intrapartum-related events (IPRE) and prematurity; among children, CSMF increases for malaria and decreases for pneumonia.

**Interpretation:** We generate VA misclassification rate estimates across two age groups, three CCVA algorithms, and eight countries. These publicly available estimates enable calibration of VA-only data from any country without needing access to CHAMPS data. The analysis also highlights systematic algorithm biases, providing direction for future improvements.

**Research in context:** **Evidence before this study:** Computer-coded verbal autopsy (CCVA) algorithms are routinely used to determine individual causes of death (COD) and population-level cause-specific mortality fractions (CSMF). However, these algorithms frequently misclassify the COD, leading to biased CSMF estimates. A recently developed framework, VA-calibration [1,2], corrects this bias by accounting for CCVA misclassification rates, estimated using COD inferred from minimally invasive tissue sampling (MITS) from the Child Health and Mortality Prevention Surveillance (CHAMPS) Network. However, there is significant variation in the VA misclassification rates across countries. Currently, VA-calibration does not account for this country-level variation, which diminishes CSMF estimation accuracy in a target population.

**Added value of this study:** We utilize the CHAMPS data and a recent Bayesian approach [3], and estimate country-specific CCVA misclassification rates. We compare the misclassification rates observed in CHAMPS with their estimates from homogeneous and county-specific models for three widely used CCVA algorithms (EAVA, InSilicoVA, InterVA), two age groups (neonates aged 0-27 days and children aged 1-59 months), and eight countries (Bangladesh, Ethiopia, Kenya, Mali, Mozambique, Sierra Leone, South Africa, and other). To demonstrate their practical application, we apply Mozambique-specific misclassification rates to VA-only data from the Countrywide Mortality Surveillance for Action (COMSA) project in Mozambique and produce national-level calibrated CMSF estimates for neonates and children.

We report three main findings. First, the country-specific estimates of the VA misclassification rates are more concordant with their observed rates in CHAMPS compared to their estimates from the homogeneous model, reducing average absolute loss by 34-38% for neonates and 13-24% for children. Second, each algorithm exhibits a systematic pattern of misclassification, consistently over- or underpredicting certain causes. Third, consistent with previous findings, calibrating COMSA data using Mozambique-specific misclassification estimates leads to notable shifts from uncalibrated CSMF estimates across the three algorithms. Among neonates, estimates generally increase for sepsis/meningitis/infection, while those for intrapartum-related events (IPRE) and prematurity decrease. The estimates among children rise for malaria and decline for pneumonia.

**Implications of all the available evidence:** We produce an inventory of VA misclassification rates resolved by two age groups, three CCVA algorithms, and country. These estimates will be made publicly available, serving as a vital resource for calibrating VA-only data from any country. The systematic biases in the algorithms quantified by the analysis provide valuable insights into the algorithms’ functioning, providing opportunities for their future improvements.

Accurate mortality data are fundamental to designing effective public health policies and achieving the Sustainable Development Goals. This research is thus highly relevant to global health, particularly for children under age five in low- and middle-income countries where vital registration systems remain incomplete. With the growing reliance on computer-coded algorithms, amid rapid AI advancements in cause-of-death determination, and their known risk of misclassification, our proposed integration of VA-calibration into the verbal autopsy workflow offers a crucial advancement in improving the accuracy of AI-powered mortality surveillance.

**Key Messages:**

- We improve VA-calibration by using CHAMPS data and a country-specific Bayesian model to account for systematic and cross-country variation in CCVA misclassification rates.
- We provide uncertainty-quantified, country-specific misclassification estimates across two age groups, three CCVA algorithms, and eight country categories (including an ‘other’ group for countries outside CHAMPS), enabling VA-calibration for any country without requiring access to CHAMPS data.
- We showcase their utility by using Mozambique-specific misclassification estimate to calibrate VA-only data from Mozambique’s COMSA project, refining CSMF estimates among neonates and children.

## 1 Introduction

Conventional cause-of-death (COD) diagnostic procedures, like full or limited autopsies, are often challenging to implement in low- and middle-income countries due to their resource-intensive nature [4]. Verbal autopsy (VA) provides a less invasive alternative for estimating COD in community settings [5–7]. VA systematically interviews caregivers using a WHO-standardized questionnaire to gather reports of the decedents’ illness signs and symptoms and available health records. The survey responses are either reviewed by physicians to assign a probable cause of death or input into computer-coded VA (CCVA) algorithms to estimate a COD (VA-COD).

Many specialized CCVA algorithms have been developed to predict COD from VA records; for example, Expert Algorithm VA (EAVA) [8], InSilicoVA [9], InterVA [10], Tariff [11], the King and Lu method [12], and domain adaptation-based methods [13,14]. The openVA R-package integrates many of these tools into one software [15,16]. Generic classifiers such as random forests [17], naive Bayes classifiers [18], and support vector machines [19] have also been employed [11,20,21]. Due to its minimally invasive nature and scalability enabled by CCVA algorithms, VA is being widely used to predict COD and build nationally representative VA-COD databases in many countries. Examples include the Countrywide Mortality Surveillance for Action (COMSA) programs in Mozambique (COMSA-Mozambique) and Sierra Leone (COMSA-SL) [22–24]. Individual-level VA data are often naively aggregated to estimate age-specific national and sub-national cause-specific mortality fractions (CSMFs), the proportion of deaths in each age group due to specific causes [25–28]. These efforts contribute to achieving health-related Sustainable Development Goals [29].

Although CCVA algorithms are essential for large-scale mortality surveillance, they often misclassify COD compared to medical certification, full autopsy, or minimally invasive tissue sampling (MITS), leading to biased CSMF estimates [1,5,30]. The VA-calibration framework addresses this by using paired VA and ‘gold standard’ (e.g., MITS) data to estimate misclassification rates and improve CSMF estimation accuracy [1,2]. This approach was recently applied in Mozambique, combining VA-only data from COMSA-Mozambique with limited MITS-VA data from the Child Health and Mortality Prevention Surveillance (CHAMPS) project [23,31,32].

To counteract limited CHAMPS data, VA-calibration originally pooled data across countries, assuming uniform misclassification rates across countries. While this improved precision, recent findings reveal significant cross-country variation, challenging that assumption [3]. To address this, Pramanik et al. [3] proposed a country-specific misclassification matrix modeling framework that models global (shared) patterns underlying misclassification, mitigates limited samples, and improves CSMF estimation accuracy.

We make two key contributions. First, we apply the framework to CHAMPS data, estimating uncertainty-quantified country-specific misclassification rates for two age groups (neonate aged 0-27 days and child aged 1-59 months) across three widely used CCVA algorithms: EAVA (deterministic), and InSilicoVA, and InterVA (both Bayesian). Second, it identifies systematic biases in each algorithm’s COD prediction, revealing previously unknown aspects of their functioning and informing future refinements. Lastly, the study demonstrates the value of the country-specific misclassification estimates by calibrating COMSA-Mozambique data, showing enhanced CSMF estimation accuracy in global mortality surveillance.

## 2 Data

### 2.1 COMSA-Mozambique VA data

The Countrywide Mortality Surveillance for Action program in Mozambique (COMSA-Mozambique) was motivated by an interest in determining CODs across all age groups using representative samples [23]. Provincially representative pregnancy and mortality data were collected through routine community surveillance in 700 clusters, each with around 300 households. Deaths were recorded at the community level, followed by interviews with caregivers using an integrated verbal and social autopsy questionnaire based on the 2016 WHO Verbal Autopsy instrument. Currently, misclassification rates for VA can only be estimated for neonates and children under five, due to the availability of CHAMPS data for these age groups. As such, our analysis is limited to these populations and includes 1,192 neonatal (0-27 days) and 2,812 child (1-59 months) records from January 2018 to December 2023.

Predicted causes of death from EAVA, InSilicoVA, and InterVA were grouped into six broad categories for neonates and nine for children. Neonatal causes included congenital malformation, pneumonia, sepsis/meningitis/infections, intrapartum-related events (IPRE), prematurity, and ‘other’. Child causes encompassed malaria, pneumonia, diarrhea, severe malnutrition, HIV/AIDS, injury, neonatal causes (IPRE, congenital malformation, prematurity), other infections, and ‘other’.

### 2.2 CHAMPS data

The Child Health and Mortality Prevention Surveillance (CHAMPS) Network collects premortem clinical and laboratory records and post-mortem VA and MITS from sites in Bangladesh, Ethiopia, Kenya, Mali, Mozambique, Sierra Leone, and South Africa. A panel of physicians and scientists uses the diagnostic test results and clinical records to ascertain a causal chain. This includes the primary or underlying, immediate, and intermediate causes leading to death. Here we only include the primary cause (*CHAMPS cause* from here on) for 1379 neonatal records and 1080 records for children (see Figures S1-S2 for neonates and Figures S12-S13 for children for a description). All deaths occurred between December 2016 and June 2023 and were grouped into the broad causes mentioned above. To analyze the accuracy of CCVA algorithms, we utilize the CHAMPS cause as the gold standard or reference COD.

## 3 Methods

### 3.1 VA-calibration and heterogeneity

For VA-only studies like COMSA-Mozambique, the *raw* or *uncalibrated estimate* of CSMF 𝑞_𝑗_ for cause 𝑗 is obtained as

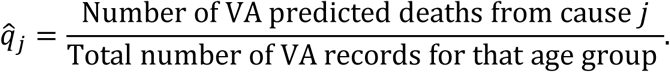

Under misclassification, they differ from their true values according to the calibration equation 𝑞_𝑗_ = ∑^𝐶^_*i*=1_ 𝜙_𝑖𝑗_𝑝_𝑖_ [1,2]. Here 𝐶 is the total number of causes, 𝚽 = (𝜙_𝑖𝑗_) is the 𝐶 × 𝐶 misclassification matrix, 𝜙_𝑖𝑗_ is the rate at which the algorithm classifies CHAMPS cause 𝑖 as cause 𝑗 (diagonals 𝜙_𝑖𝑖_’s are sensitivities, and off-diagonals 𝜙_𝑖𝑗_’s are false negatives), and 𝑝_𝑖_ is the true CSMF of cause 𝑖 (CHAMPS cause is assumed as the truth). While uncalibrated estimates appear more precise, they ignore misclassification as evidenced in CHAMPS, risking biased and overconfident results. Leveraging limited paired CHAMPS-VA COD data, VA-calibration solves an inverse problem to correct for this bias. It produces a *calibrated CSMF estimate* which increases uncertainty but improves out-of-sample predictive performance (see [1,2] and Section 4 in [3]).

Conducting MITS and obtaining CHAMPS causes are time-sensitive and resource-intensive, making them mostly available in hospital settings and resulting in limited samples. To overcome this, the current VA-calibration approach pools CHAMPS data across countries to improve sample size and precision in estimating misclassification rates.

Pooling assumes the same VA misclassification across countries, but its effectiveness relies on how well they represent the study country [2]. While this assumption cannot be tested in VA-only settings due to unavailable CHAMPS-CODs, it rests on the idea of globally similar symptom-cause relationships. In contrast, CHAMPS data show significant cross-country variation in misclassification rates (e.g., see Figure 1 for EAVA and Supplement Figure S3), potentially biasing CSMF estimates.

**Figure 1.**
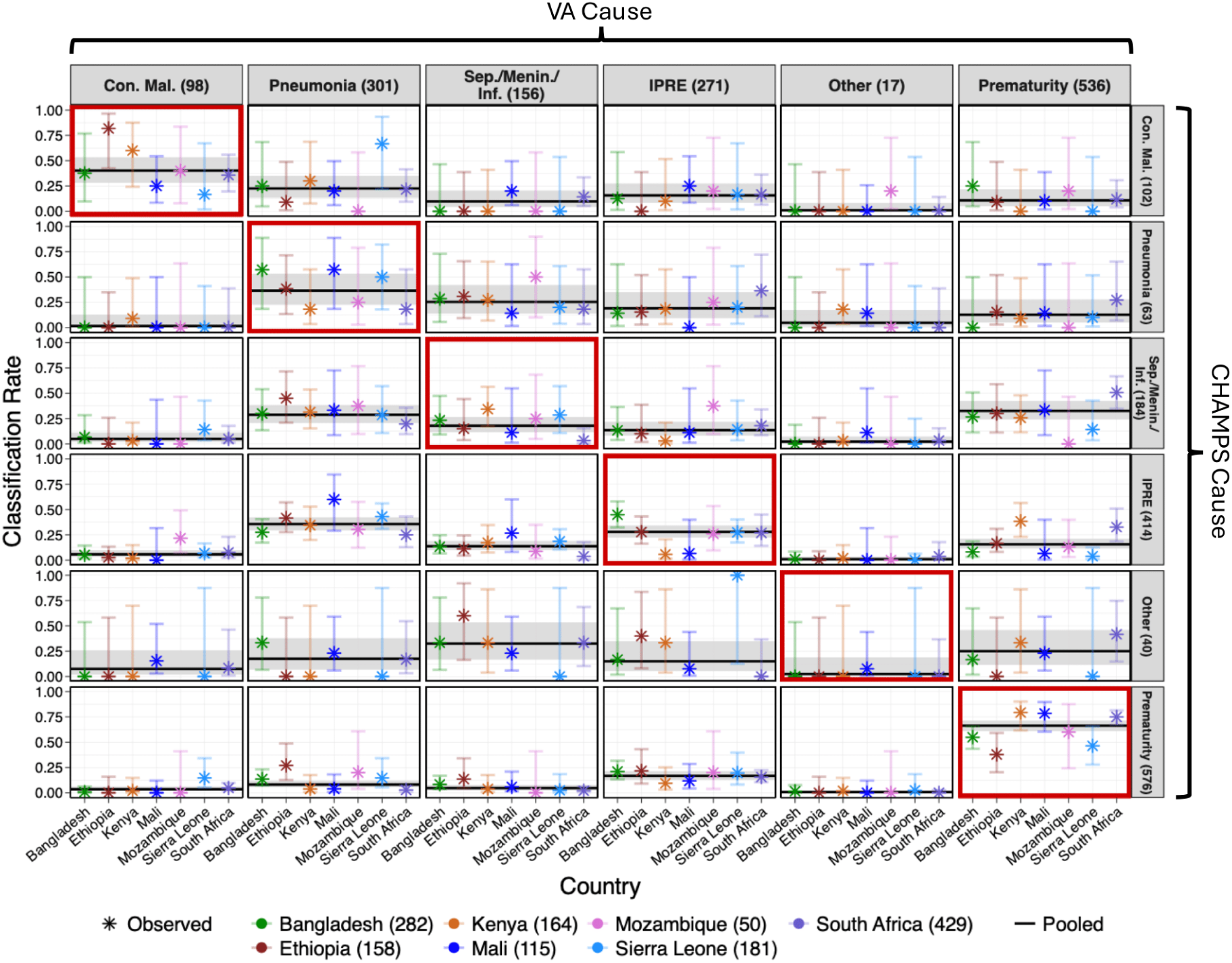
Observed misclassification rates of EAVA for neonates (0-27 days) in CHAMPS. Rows and columns indicate CHAMPS and VA causes, with combined sample sizes across countries indicated in parentheses. Sensitivities and false negatives are respectively along the diagonal (red) and off-diagonal panels. Misclassification rates are conditioned on CHAMPS cause (row), so values in each row sum to 1 for each country. Con. mal., Sep./Menin./Inf., IPRE respectively denote congenital malformation, sepsis/meningitis/infection, and intrapartum-related events.

### 3.2 Modeling Structure and Heterogeneity in VA Misclassification

Pramanik et al. [3] proposed an efficient Bayesian modeling framework that improves precision in country-specific VA misclassification estimation under limited samples (see Supplement Sections S1.1-S1.4). The model incorporates the following key:

1. They introduce a parsimonious base model based on two underlying latent mechanisms, *intrinsic accuracy* and *pull*, that characterize global misclassification patterns. Intrinsic accuracy reflects the algorithm’s ability by design to correctly identify a true cause. Pull captures systematic bias, indicating an algorithm’s tendency to over- or underpredict certain causes regardless of the true cause when it fails to correctly identify the true cause by design. Together, they promote model parsimony, improving efficiency under limited. The framework builds on this and extends to country-specific modeling.
2. The framework adaptively chooses its complexity based on the data using continuous shrinkage. This balances the bias-variance tradeoff, favoring simpler models under limited samples or the absence of evidence.
3. The model produces uncertainty-quantified, country-specific misclassification estimates for seven CHAMPS countries, along with an estimate for countries outside CHAMPS. The estimate for other countries is centered on the pooled rate, with uncertainty reflecting the degree of homogeneity within CHAMPS. This enables VA-calibration in any country without requiring direct access to CHAMPS data. Additionally, estimates of intrinsic accuracy and pull reveal new insights into how CCVA algorithms function.

The framework improves VA misclassification estimates, leading to more accurate CSMF estimates when calibrating VA-only data like COMSA-Mozambique.

### 3.3 Modular VA-calibration Using Country-Specific VA Misclassification Algorithm-specific calibration

The current VA-calibration approach jointly models CHAMPS and VA-only data, requiring access to both. [31]. We apply the modular VA-calibration from [3]. This analyzes the CHAMPS data once using the framework in [3] and stores uncertainty-quantified estimates of misclassification matrices, resolved by country, age group, and CCVA algorithm. To calibrate a VA-only study like COMSA-Mozambique, country-, age-, and algorithm-specific misclassification estimates are used as informative priors, enabling calibration without direct access to CHAMPS data (see Section S1.6 in the supplement and 3.8 in [3]). Figure 2 outlines the calibration pipeline. We exclude ‘other’ as it comprises different causes in VA and CHAMPS diagnoses, and misclassification matrices are renormalized to remain valid.

**Figure 2.**
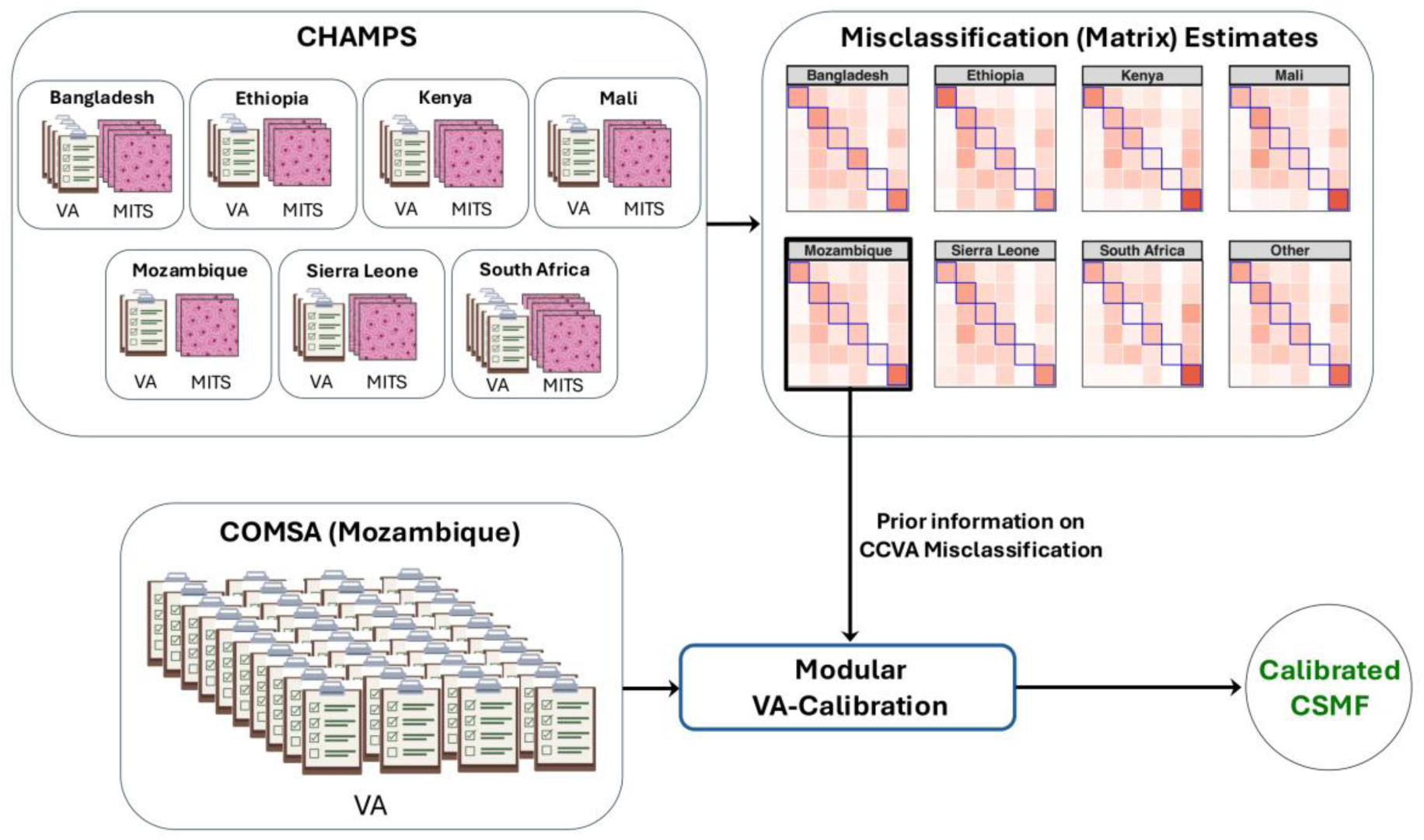
The modular VA-Calibration pipeline for generating calibrated verbal autopsy (VA)-based cause-specific mortality fractions (CSMF) in the Countrywide Mortality Surveillance for Action (COMSA) in Mozambique. Country-specific misclassification estimates at the top are obtained using limited paired cause of death (COD) diagnoses from minimally invasive tissue sampling (MITS) and VA collected through the Child Health and Mortality Prevention Surveillance (CHAMPS) project.

#### Ensemble calibration

CCVA algorithms often disagree in predicting COD, making it challenging to identify the most accurate algorithm. Following [1,2], we perform ensemble VA-calibration by integrating misclassification rates of all algorithms in a Bayesian model. This lowers the risk of using an inaccurate algorithm and improves CSMF estimates.

## 4 Results

We present VA misclassification analysis using CHAMPS data and apply Mozambique-specific estimates to COMSA-Mozambique. Results for children are summarized briefly, with full details in Section S3 of the supplement.

### 4.1 VA Misclassification Estimates from CHAMPS Analysis

#### 4.1.1 Neonatal Deaths (0-27 Days)

##### VA misclassification rates and model comparison

Figure 3 compares observed misclassification rates with estimates from the homogeneous (left) and country-specific (right) models for the three algorithms. Two key insights emerge: first, the country-specific estimates align more closely with the observed rates, as seen by their proximity to the 𝑦 = 𝑥 line. Second, in the country-specific model, larger points (more samples) are closer to the 𝑦 = 𝑥 line, reflecting the model’s greater reliance on observed rates when sample sizes are sufficient. The few points that deviate from the 𝑦 = 𝑥 line in the country-specific model reflect low sample sizes. In such cases, the model shrinks toward the pooled estimate, causing deviations from observed rates. To quantify the improvement, we calculate the average absolute difference between the modeled estimate 𝚽̃_𝑠_ = (𝜙̃_𝑠𝑖𝑗_) and the observed rate 𝚽^_𝑠_ = (𝜙^_𝑠𝑖𝑗_) across all countries and cause pairs, using the absolute loss function ∑_𝑠𝑖𝑗_ |𝜙̃_𝑠𝑖𝑗_ − 𝜙^_𝑠𝑖𝑗_|⁄𝑂𝑏𝑠, where 𝑂𝑏𝑠 is the total number of observed CHAMPS-VA cause pairs. Supplementary Figures S3-S4 and S6-S8 compare observed and estimated rates for both models. Figure S11 presents effect size estimates highlighting evidence of systematic pull and cross-country heterogeneity.

**Figure 3.**
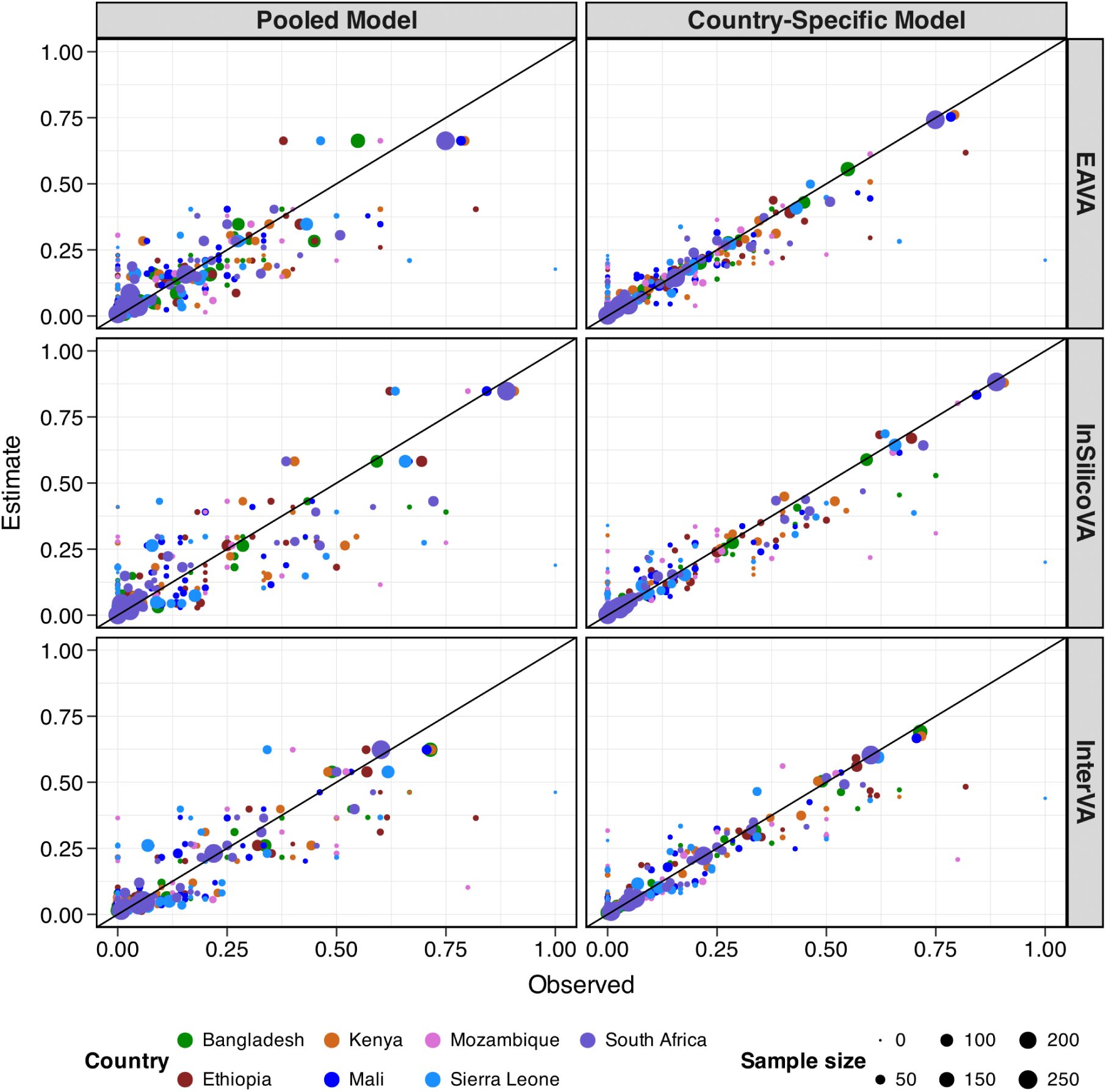
Scatterplots of observed (x-axis) and estimated (y-axis) misclassification rates of EAVA (top row), InSilicoVA (middle row), and InterVA (bottom row) for neonatal deaths (0-27 days) in CHAMPS. It compares estimates from the homogeneous or pooled model (left panels) and the country-specific model (right panels). Countries are denoted in different colors. The point sizes reflect the observed sample size for each corresponding CHAMPS cause. The black line corresponds to the 𝑦 = 𝑥 line. Compared to the homogeneous model, the country-specific model reduces the average absolute loss with respect to observed rates by 35%, 38%, and 34% for the three algorithms for this age group.

Figure S4 in the supplement presents the estimated country-specific misclassification rates. The leftmost column shows rates pooled across countries, highlighting substantial variation in sensitivity by cause and across CCVA algorithms. Sensitivity is highest for prematurity (ranging from 62% to 85%) and lowest for sepsis/meningitis/infection (12-22%) and ‘other’ (5-10%). Algorithm performance also varies: InSilicoVA has difficulty detecting congenital malformations, while EAVA and InterVA achieve sensitivities of about 41% and 35%, respectively. Pooled false negative rates are notable as well; for example, 31-44% of sepsis/meningitis/infection deaths are misclassified as prematurity, and 12-39% of congenital malformation deaths are misclassified the same way, depending on the algorithm.

Relative to the homogeneous model, the country-specific model improved point estimates for 72-91% of cause pairs, reducing 44-50% absolute bias on average. For EAVA, InSilicoVA, and InterVA, the country-specific model reduced average absolute loss by 35%, 38%, and 34%, respectively (see Table 1). Additionally, it enhanced uncertainty quantification for 62-70% of cause pairs, with 75-79% reductions on average in interval scores (see supplemental Figures S9-S10).

**Table 1.** Improvement in misclassification estimates for neonates (0-27 days) when using a country-specific model compared to a homogeneous (pooled) model. For each model, we compute the average absolute difference ∑_𝑠𝑖𝑗_ |𝜙̃_𝑠𝑖𝑗_ − 𝜙^_𝑠𝑖𝑗_|⁄𝑂𝑏𝑠 (the absolute loss function) between the modeled estimate 𝜱̃_𝑠_ = (𝜙̃_𝑠𝑖𝑗_) and their observed rate 𝜱^_𝑠_ = (𝜙^_𝑠𝑖𝑗_) in the country 𝑠 and 𝑂𝑏𝑠 is the total number of observed CHAMPS-VA cause pairs and countries. The table reports the percentage improvement in loss by country-specific model over the homogeneous model.

| **EAVA** | **InSilicoVA** | **InterVA** |
| --- | --- | --- |
| 35 | 38 | 34 |

##### Estimate of intrinsic accuracy and pull

Figure 4 illustrates estimates of intrinsic accuracy and pull, and serves as diagnostics of CCVA algorithms. All algorithms have the highest intrinsic accuracy for prematurity, with InSilicoVA performing best for this cause. The accuracy is the lowest for ‘other’. Regarding pull, no preference for an algorithm indicates a uniform pull of 1/6 for six causes, with deviations from 1/6 suggesting a systematic bias. EAVA favors pneumonia but underpredicts congenital and ‘other’; InSilicoVA overpredicts prematurity and underpredicts congenital and ‘other’; and InterVA favors IPRE and prematurity while down-weighting other causes. These diagnostics reveal algorithm-specific tendencies.

**Figure 4.**
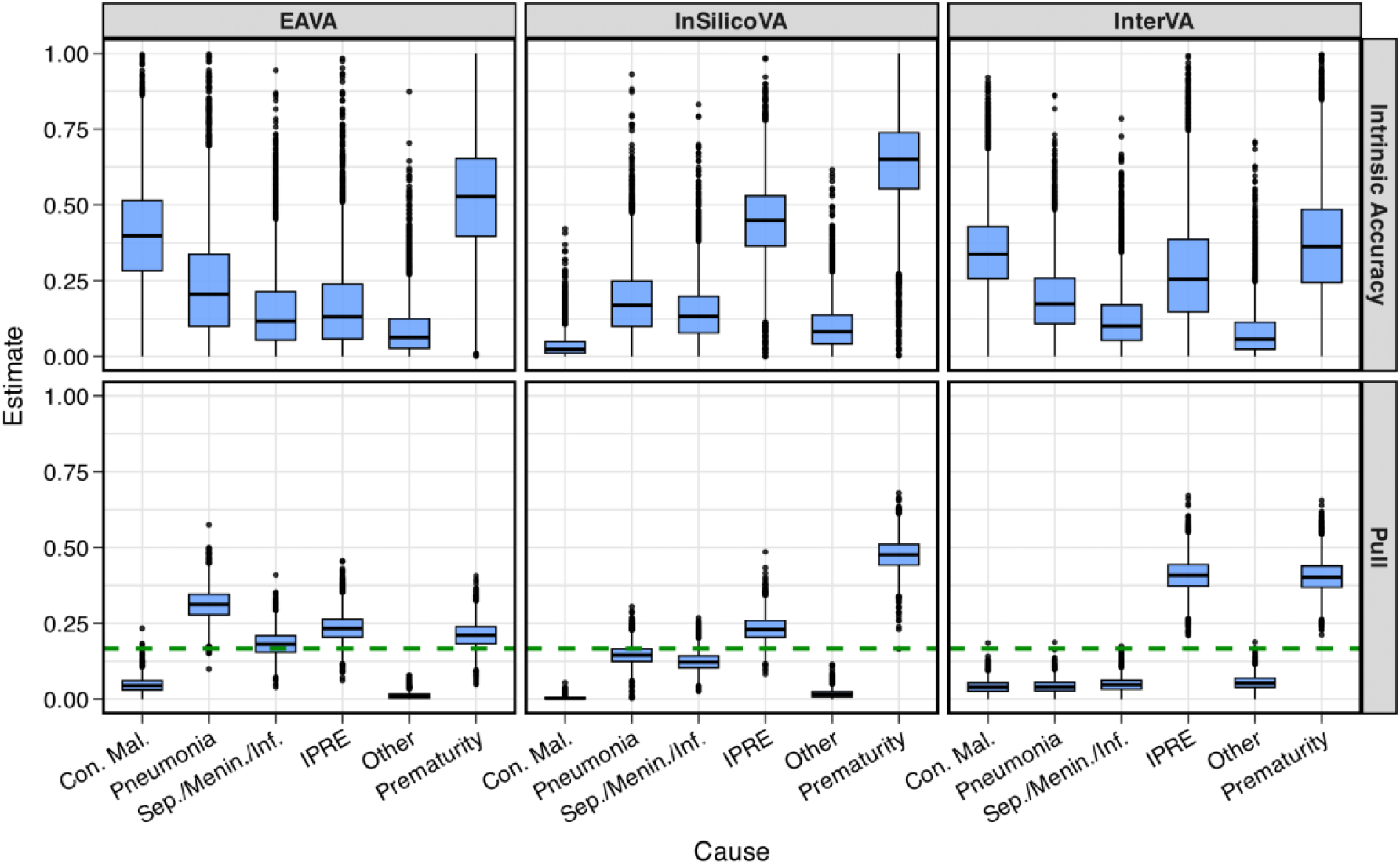
Posterior estimates of intrinsic accuracy (top) and pull (bottom) for EAVA, InSilicoVA, InterVA algorithms in CHAMPS among neonatal deaths (0-27 days). Bottom panel: For reference, the green horizontal dashed line indicates no preference, corresponding to a uniform pull of 1/6 for each of the six causes. Con. mal., Sep./Menin./Inf., IPRE denote congenital malformation, sepsis/meningitis/infection, and intrapartum-related events.

##### Examining the sources of heterogeneity in VA misclassification

The left panel in Figure 5 highlights variability in EAVA’s sensitivity for sepsis/meningitis/infection, which is high in Kenya (31%) but very low in South Africa (6%). This is partly due to EAVA’s hierarchical structure, where sepsis ranks low, only above jaundice, neonatal hemorrhage, and sudden unexplained infant death, which are categorized as ‘other’. Among 61 deaths with CHAMPS cause sepsis/meningitis/infection, 37 met clinical criteria for sepsis by EAVA and only two were assigned sepsis. The other 35 records have an EAVA cause, in addition to sepsis, which is a higher-up in the hierarchy: 2 congenital malformations, 8 intrapartum-related events, 12 pneumonia, and 13 prematurity. This highlights the need for multi-cause analysis to improve the accuracy of VA.

**Figure 5.**
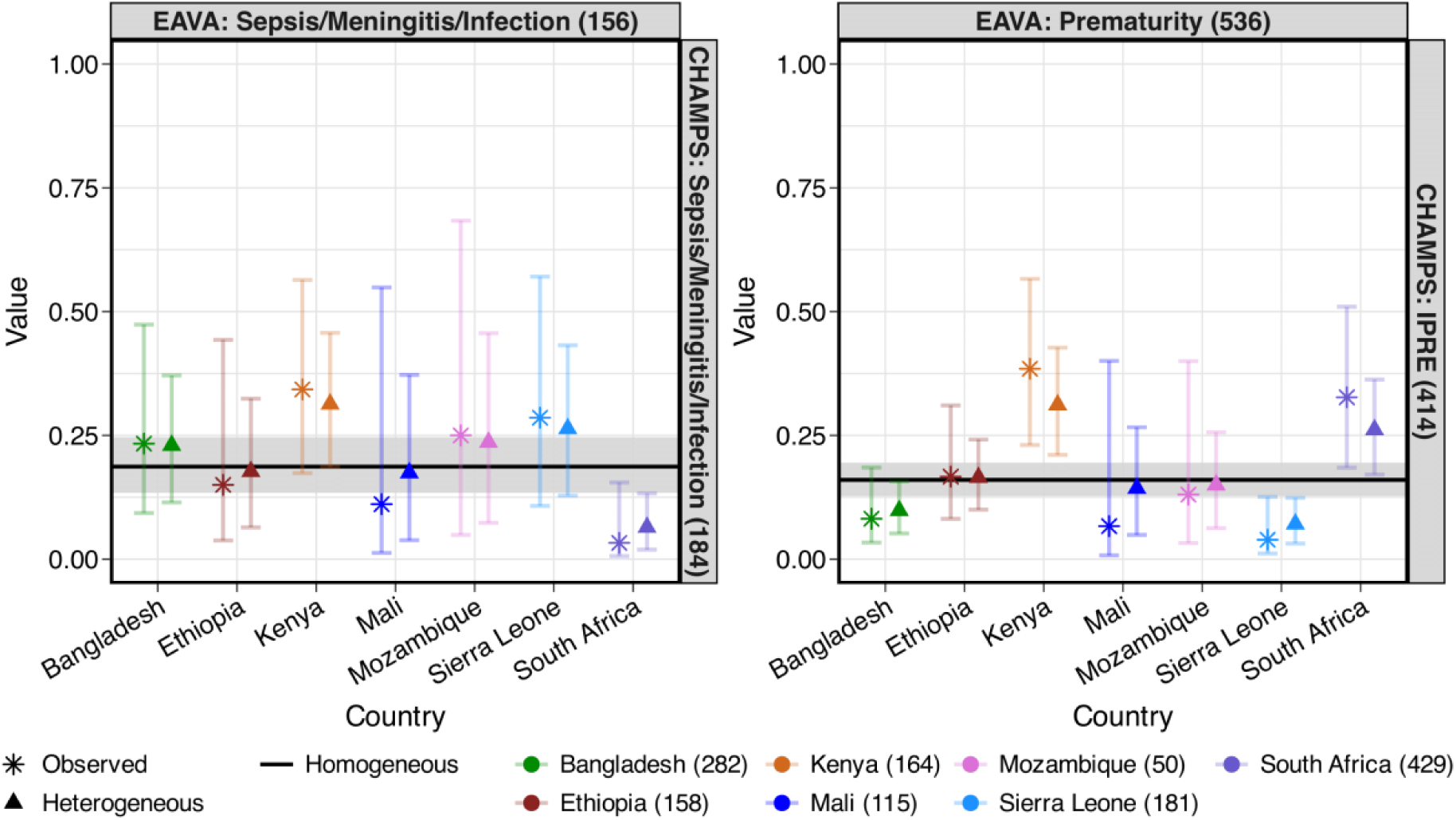
Evidence for cross-country heterogeneity for EAVA misclassification among neonatal deaths (0-27 days). Left panel: Observed and estimated sensitivities for sepsis/meningitis/infection. Right panel: Observed and estimated false negative rates for CHAMPS cause IPRE and VA cause prematurity. IPRE denotes intrapartum-related events.

The right panel of Figure 5 shows higher false-negative rates for the CHAMPS-VA cause pair IPRE-prematurity in Kenya and South Africa, and all records meet EAVA’s prematurity criteria. IPRE (due to birth injury or asphyxia) ranks top third in the hierarchy above prematurity.

Diagnosing birth asphyxia requires answering VA question id10106 (“How many minutes after birth did the baby first cry?”), which is missing in 7 of 20 deaths in Kenya and 11 of 17 deaths in South Africa. This highlights the need to improve data quality through better tools and interviewer training.

#### 4.1.2 Summary of VA misclassification rates for children (1-59 months)

Analysis of VA misclassification for child deaths in CHAMPS shows significant variability across causes, algorithms, and countries (Figures S14-S19 in the supplement). Overall, InSilicoVA and InterVA have very low sensitivity for neonatal causes (3-4%), while EAVA’s sensitivity is higher at 37%. False negative rates are substantial, especially for malaria and ‘other infection’ (25-30%), with notable country-level variation; for example, EAVA’s sensitivity for severe malnutrition ranges from 9-28%, and false negatives for malaria-diarrhea vary between 7-18%.

The country-specific misclassification model outperforms the homogeneous model by reducing absolute bias in 69-72% of cause pairs with 27-40% bias reduction on average, and improving uncertainty quantification with 56-68% lower interval scores in 65-69% of cause pairs. For children, it also lowers average absolute loss by 19%, 24%, and 13% for EAVA, InSilicoVA, and InterVA (Table 2). Detailed results are in Section S3.1.

**Table 2.** Improvement in misclassification estimates for children (1-59 months) when using a country-specific model compared to a homogeneous (pooled) model. For each model, we compute the average absolute difference ∑_𝑠𝑖𝑗_ |𝜙̃_𝑠𝑖𝑗_ − 𝜙^_𝑠𝑖𝑗_|⁄𝑂𝑏𝑠 (the absolute loss function) between the modeled estimate 𝜱̃_𝑠_ = (𝜙̃_𝑠𝑖𝑗_) and their observed rate 𝜱^_𝑠_ = (𝜙^_𝑠𝑖𝑗_) in the country 𝑠 and 𝑂𝑏𝑠 is the total number of observed CHAMPS-VA cause pairs and countries. The table reports the percentage improvement in loss by country-specific model over the homogeneous model.

| **EAVA** | **InSilicoVA** | **InterVA** |
| --- | --- | --- |
| 19 | 24 | 13 |

### 4.2 Case Study: CSMF Estimates in Mozambique Using COMSA-Mozambique

#### 4.2.1 Neonates (0-27 Days)

##### Misclassification estimates from CHAMPS as prior information

Figure 6 presents the (expected) Mozambique-specific misclassification estimates from analyzing CHAMPS data. They are utilized as an informative prior for modular VA-calibration of VA-only COD data from COMSA-Mozambique. Sensitivities are generally high for prematurity (particularly in InSilicoVA) and IPRE (except EAVA). False negatives are evident across all algorithms, especially for VA causes IPRE and prematurity.

**Figure 6.**
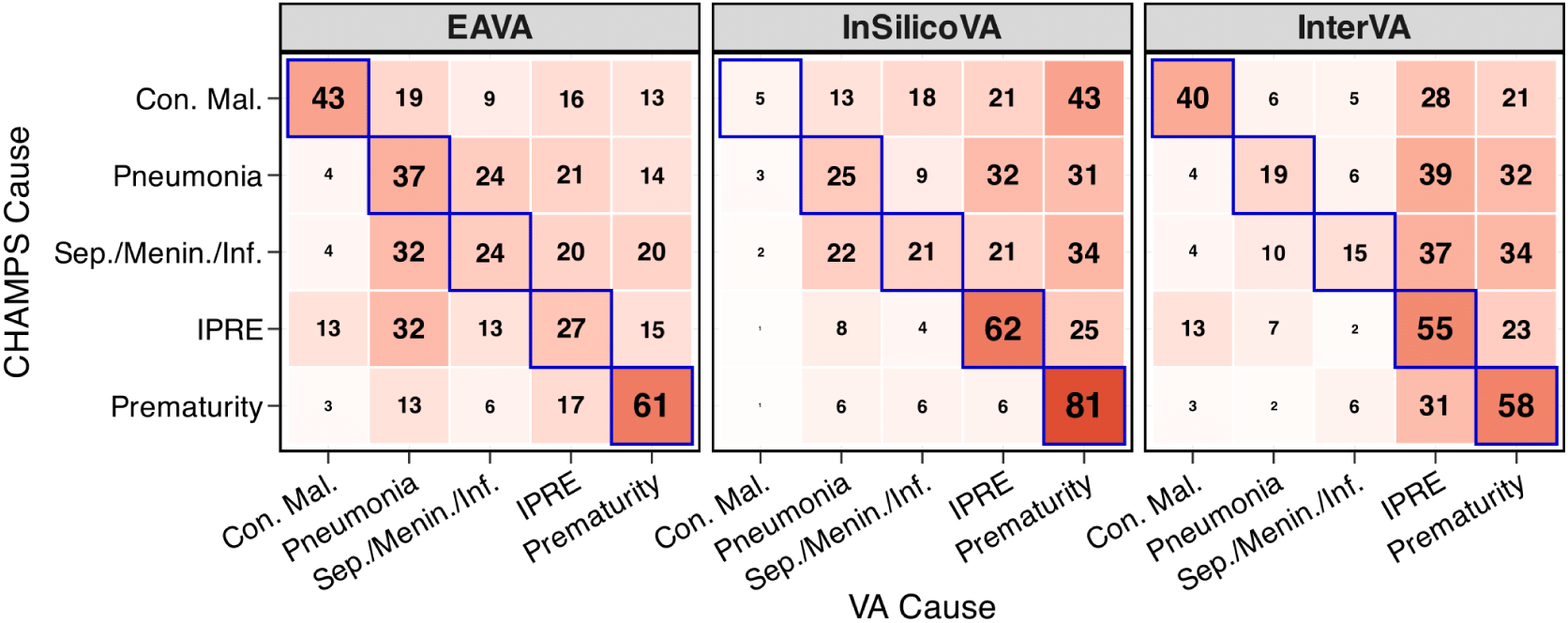
Obtained from CHAMPS analysis of neonates (0-27 days) deaths, this is the (expected) misclassification (without ‘other’) for Mozambique that is used as an informative prior in the modular VA-calibration. Sensitivities are along diagonals (outlined in blue). Con. mal., Sep./Menin./Inf., IPRE denote congenital malformation, sepsis/meningitis/infection, and intrapartum-related events.

##### Raw CSMF estimates

Figure 7 presents raw neonatal CSMF estimates (blue) in Mozambique from VA-only data. Among 1,192 deaths, EAVA and InSilicoVA identify sepsis/meningitis/infection as the leading cause, while InterVA and the ensemble rank prematurity highest. All algorithms consistently list IPRE as the second most common cause and congenital malformations as the least common.

**Figure 7.**
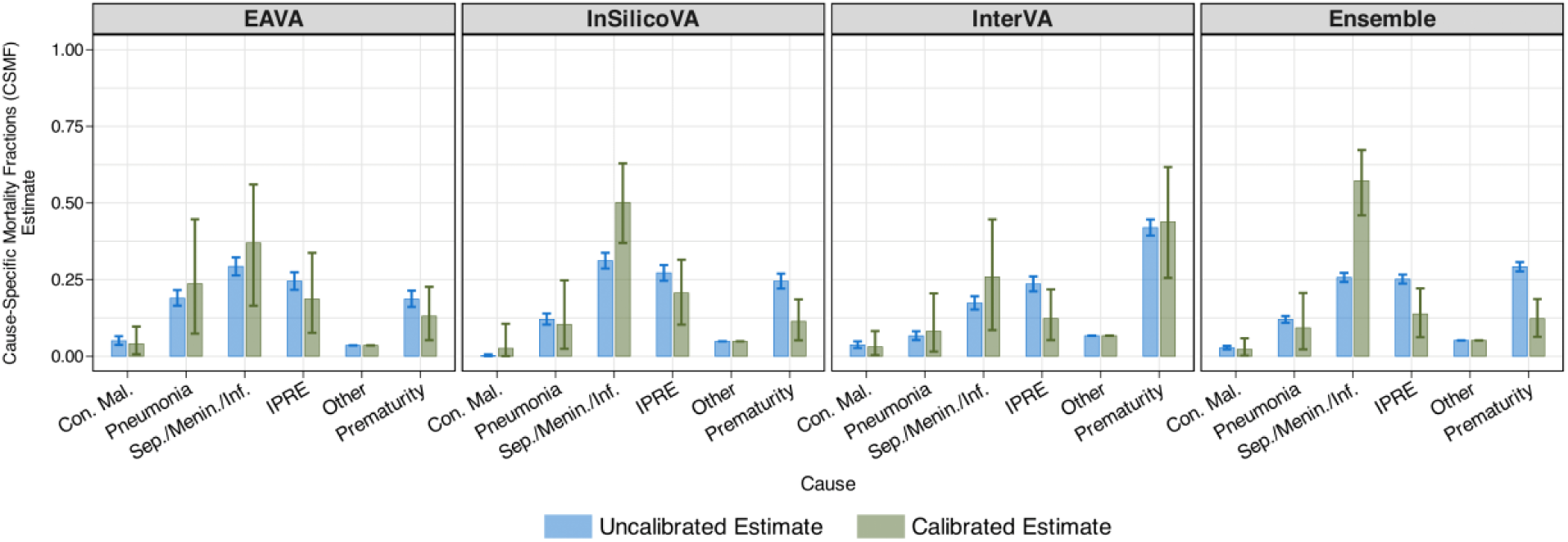
Comparison of uncalibrated (blue) and calibrated (green) Cause-Specific Mortality Fraction (CSMF) estimates for neonates (0-27 days) in Mozambique using EAVA, InSilicoVA, InterVA, and their ensemble. Bar heights represent the point estimates (posterior means), while the error bars indicate uncertainty (95% credible intervals). Although uncalibrated CSMF estimates have lower uncertainty than calibrated ones, they assume perfect classification—an assumption contradicted by substantial VA misclassification observed in CHAMPS. No calibration can lead to overconfidence and biased CSMF estimates. Con. mal., Sep./Menin./Inf., IPRE denote congenital malformation, sepsis/meningitis/infection, and intrapartum-related events.

##### Calibrated CSMF estimates

Figure 7 also shows calibrated, uncertainty-quantified CSMF estimates (green). Calibration increases the CSMF for sepsis/meningitis/infection and decreases it for IPRE and prematurity (except for InterVA). These reflect the misclassification, where the algorithms misclassify 20%, 21%, and 37% of sepsis/meningitis/infection deaths as IPRE, and 20%, 34%, and 34% of sepsis/meningitis/infection deaths as prematurity (see Figure 6). The calibration adjusts for this undercounting of sepsis/meningitis/infection deaths and overcounting of IPRE and prematurity.

The far-right panel of Figure 7 shows ensemble CSMF estimates, with calibration attributing 58% of neonatal deaths to sepsis/meningitis/infection, followed by 13% to IPRE, 12% to prematurity, and smaller shares to other causes. The 95% credible intervals confirm significant increases in sepsis/meningitis/infection (47-68% vs. 26% uncalibrated) and decreases in prematurity and IPRE (6-18% and 6-22% vs. 29% and 25% uncalibrated). For other causes, calibrated intervals overlap with uncalibrated values. These results align with previous findings in [31].

#### 4.2.2 Children (1-59 Months)

##### Misclassification estimates from CHAMPS

For the three CCVA algorithms, Figure 8 shows (expected) Mozambique-specific misclassification estimates from analyzing CHAMPS data. Sensitivity is the highest for injury and diarrhea, with high false negatives for VA causes pneumonia, diarrhea, and ‘other infections’.

**Figure 8.**
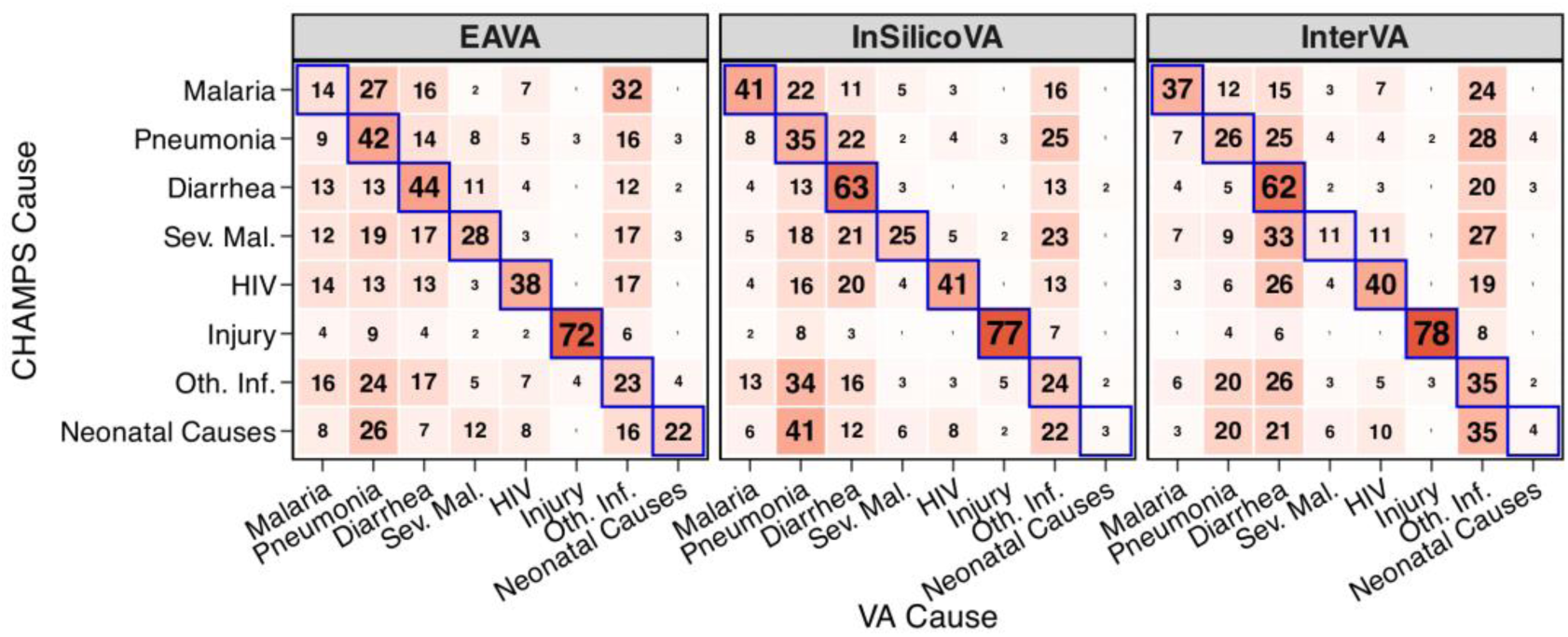
Obtained from CHAMPS analysis of child (1-59 months) deaths, this is the (expected) misclassification (without ‘other’) for Mozambique that is used as an informative prior in the modular VA-calibration. Sensitivities are along diagonals (outlined in blue). Sev. mal. and Oth. Inf. denote severe malnutrition and other infections

##### CSMF estimates

Figure 9 compares uncalibrated (blue) with calibrated (green) CSMF estimates for 2,812 VA-only child deaths from COMSA in Mozambique. Calibration generally increases the estimated CSMF for malaria and decreases it for pneumonia and diarrhea. The 95% credible intervals typically include the uncalibrated estimates but lie near the edges for some causes, especially ‘other infections’ (EAVA), malaria (InterVA and ensemble), and pneumonia (ensemble). Overall, the findings, particularly for malaria and pneumonia, align with results from prior research [31].

**Figure 9.**
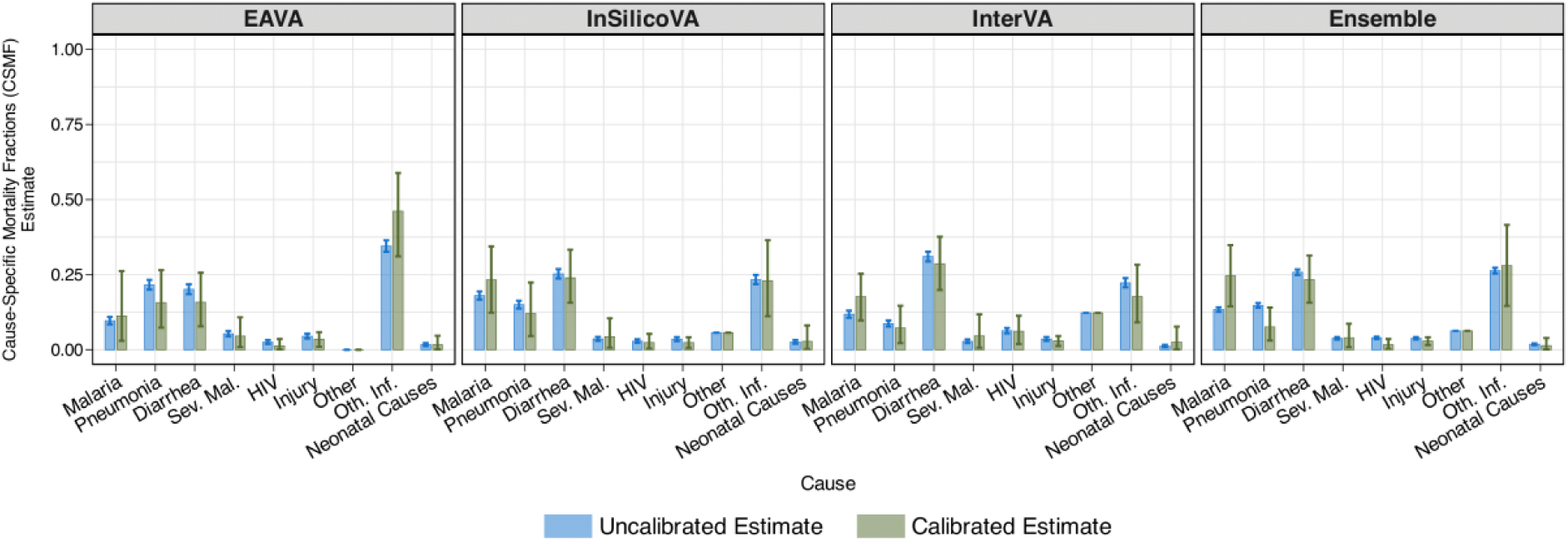
Comparison of uncalibrated (blue) and calibrated (green) Cause-Specific Mortality Fraction (CSMF) estimates for children (1-59 months) using EAVA, InSilicoVA, InterVA, and their ensemble. Bar heights represent the point estimates (posterior means), while the error bars indicate uncertainty (95% credible intervals). Although uncalibrated CSMF estimates show lower uncertainty than calibrated ones, they assume perfect classification—an assumption contradicted by substantial VA misclassification observed in CHAMPS. No calibration can lead to overconfidence and biased CSMF estimates. Sev. mal. and Oth. Inf. denote severe malnutrition and other infections.

In Tables S1 and S2 in the supplement, we include Mozambique’s age-group- and algorithm-specific CSMF estimates featured in Figure 7 and Figure 9.

## 5 Discussion

The widespread use of CCVA algorithms in VA-based mortality surveillance underscores the need for accurate misclassification rate estimates to ensure reliable CSMF estimates. Leveraging limited paired CHAMPS-VA data, covering children under five across seven countries, we highlight significant cross-country variation and systematic biases in three widely used CCVA algorithms (EAVA, InSilicoVA, and InterVA). We produce uncertainty-quantified misclassification matrix estimates by algorithm, age group (neonates and children), and country (seven CHAMPS countries plus an ‘other’ category). They will be publicly available and regularly updated, enabling calibration of VA-only data globally without direct access to CHAMPS data.

The effectiveness of VA calibration relies on the *transportability assumption* that misclassification patterns in labeled data (e.g., CHAMPS) apply to unlabeled data (e.g., COMSA-Mozambique). While this cannot be directly verified due to the absence of reference causes in the unlabeled data, it is founded on globally similar VA symptom profiles given a CHAMPS cause. The analysis also assumes that misclassification patterns are stable over time. However, real-world changes in disease patterns, healthcare access, or VA implementation challenge the assumption, making the exploration of time-varying misclassification an important direction for future research.

Misclassification rates were calculated by comparing VA-assigned causes to the underlying causes identified through CHAMPS’ Determination of Cause of Death (DeCoDe) process, consistent with prior findings that VA typically targets the underlying cause [33]. Given the high observed misclassification, further research is needed to assess whether VA may instead capture a different cause within the broader causal chain, suggesting a multi-cause framework.

## Data Availability

All data produced in this research are available upon request to the authors and will be made publicly available soon.

## Acknowledgement

S.P., E.W., H.K., S.Z., and A.D. were supported by the Bill and Melinda Gates Foundation Grant INV-034842.

## Competing interests

The authors declare that they have no competing interests.

## Ethical Approval

The research does not qualify as human subject research. We use the anonymized verbal autopsy cause of death data collected in the Comprehensive Mortality Surveillance for Action (COMSA) program in Mozambique (COMSA-Mozambique; OPP1163221; PI: Drs. Amouzou and Macicame). We are using unidentifiable cause of death data that are collected by the Child Health and Mortality Prevention Surveillance (CHAMPS) Network. This is considered non-human subjects research under HHS human subject regulation (45 CFR Part 46): “*(4) Secondary research for which consent is not required: Secondary research uses of identifiable private information or identifiable biospecimens, if at least one of the following criteria is met: (i) The identifiable private information or identifiable biospecimens are publicly available; (ii) Information, which may include information about biospecimens, is recorded by the investigator in such a manner that the identity of the human subjects cannot readily be ascertained directly or through identifiers linked to the subjects, the investigator does not contact the subjects, and the investigator will not re-identify subjects;*”. The data used here are anonymized in such a way that the identity of the human subjects cannot readily be ascertained directly or through identifiers linked to the subjects. This research uses the data already collected in COMSA-Mozambique and CHAMPS and only performs statistical analyses with them.

The ethical approval for CHAMPS was obtained from the ethical review boards at each participating site, as well as from the Emory University Rollins School of Public Health (Emory Institutional Review Board: 00091706) and the U.S. Centers for Disease Control and Prevention (CDC). Written informed consent was requested and obtained from the primary caregivers of all reported child deaths within the CHAMPS catchment areas before the collection of data and specimens. Families were provided with the results regarding the cause of death of their deceased child.

COMSA-Mozambique received ethical approval from the National Health Bioethics Committee of Mozambique (REF 608/CNBS/17) and the Institutional Review Board of the Johns Hopkins Bloomberg School of Public Health (IRB#7867).

## Notes

### Competing Interest Statement

The authors have declared no competing interest.

### Author Declarations

CHAMPS: The ethical approval was obtained from the ethical review boards at each participating site, as well as from the Emory University Rollins School of Public Health (Emory Institutional Review Board: 00091706) and the U.S. Centers for Disease Control and Prevention (CDC). Written informed consent was requested and obtained from the primary caregivers of all reported child deaths within the CHAMPS catchment areas before the collection of data and specimens. Families were provided with the results regarding the cause of death of their deceased child. COMSA-Mozambique: The project received ethical approval from the National Health Bioethics Committee of Mozambique (REF 608/CNBS/17) and the Institutional Review Board of the Johns Hopkins Bloomberg School of Public Health (IRB#7867).

### Summary of Updates

We have added Section S4 in the supplement. It presents uncertainty-quantified estimates of uncalibrated and calibrated cause-specific mortality fractions (CSMF) for Mozambique.

