## Supplementary Material for "Country-Specific Estimates of Misclassification Rates of Computer-Coded Verbal Autopsy Algorithms"

#### Abstract

In this supplementary file, we provide additional materials for the main article.

Disclaimer: The findings and conclusions in this report are those of the author(s) and do not necessarily represent the views of the U.S. Centers for Disease Control and Prevention.

### S1. A Brief Overview of Country-Specific Misclassification Matrix Modeling and Modular VA-Calibration

In recent work, [1] proposed a comprehensive framework for modeling misclassification matrices for computer-coded verbal autopsy (CCVA) algorithms, particularly suited for contexts with limited labeled data. Their method introduces a novel, parsimonious model for estimating a homogeneous misclassification matrix in small-sample settings, which is then expanded into a hierarchical structure allowing for increasingly flexible parameterizations. This framework accounts for cross-country variation and utilizes data-driven shrinkage techniques to favor simpler models in the absence of sample size or evidence. Additionally, they proposed a modular VA-Calibration approach that leverages uncertainty-aware misclassification matrix estimates, derived from limited labeled CHAMPS-VA cause of death (COD) data, to improve the accuracy of cause-specific mortality fractions (CSMF) using only verbal autopsy COD data from the target country. Below, we outline the key components of the two frameworks.

#### S1.1 Base Model: Parsimonious Homogeneous Modeling of Misclassification Matrices

Let $C$ is the total number of causes, $\mathbf{\Phi} = (\phi_{ij})$ is the $C \times C$ misclassification matrix, $\phi_{ij}$ is the rate at which the algorithm classifies cause of death (COD) as $j$ when the CHAMPS cause is $i$ (diagonals $\phi_{ii} = P(V = i|M = i)$ are sensitivities, and off diagonals $\phi_{ij} = P(V = j|M = i)$ are false negatives). [1] proposes a parsimonious *base model* for $\mathbf{\Phi}$, capturing two core behavioral aspects of any general algorithm: its ability *by design* to correctly identify the true CHAMPS cause, and its *systematic preference* toward predicting certain causes when it fails to identify the true CHAMPS cause.

**Intrinsic Accuracy (Diagonal Effect):** VA algorithms are primarily designed to determine the causes of death accurately. Let $a_i$ denote the *intrinsic accuracy* of an algorithm for cause $i$—the probability that it correctly assigns cause $i$ by design when that is the true cause. Conceptually, the intrinsic accuracy for predicting cause $i$ arises from a distinct pattern of symptoms in the VA record that appears only when $i$ is the true cause.

**Systematic Preference or Pull (Column Effect):** In cases where an algorithm fails to correctly identify the true cause by design, the base model assumes a simplified scenario where the algorithm assigns one of the $C$ causes independent of the true cause (for example, CHAMPS cause) with prespecified probabilities. This reflects the algorithm's *systematic preference* or *pull* toward certain causes. The pull toward cause $j$ is denoted by $\rho_j$. It represents the probability of assigning the cause $j$ irrespective of the true cause when the algorithm, by design, fails to correctly identify the true cause.

Intrinsic accuracy and pull capture key algorithmic behaviors that jointly determine the misclassification matrix. While these processes are not directly observable, the misclassification rates that we observe result from their combined effects. In the base model, a correct classification for true cause $i$ can occur in two ways: by design with probability $a_i$, or—if the designed match fails—with probability $(1 - a_i)\rho_i$ due to the algorithm's pull toward cause $i$. Conversely, a misclassification into cause $i \neq j$ happens only if the algorithm fails to match by design and is then influenced by the pull toward cause $j$, occurring with probability $(1 - a_i)\rho_j$. Under the base model, sensitivities and false negatives are thus equal to

$$\phi_{ii} = a_i + (1 - a_i)\rho_i \text{ and } \phi_{ij} = (1 - a_i)\rho_j.$$

This base model offers several advantages for modeling misclassification. It provides a parsimonious framework for representing homogeneous errors using only $2C - 1$ parameters, $C$

intrinsic accuracies and $C-1$ pull parameters, compared to $C^2-C$ free parameters in a general unstructured misclassification matrix. This reduction in dimension is especially valuable for modeling country-specific effects with limited data. Moreover, modeling pull enables us to quantify the systematic bias of an algorithm (see Figure 3 in the main article for neonates (0-27 days) and Figure S23 below for children (1-59 months)). Beyond providing more accurate estimates of misclassification rates, this approach helps identify causes that the algorithm systematically favors, offering valuable insights into its behavior and guiding improvements in CCVA algorithm design. While intrinsic accuracy and pull are not directly observable, Theorem 3.1 in [1] provides a characterization of the base model in terms of constant misclassification odds. When a VA algorithm misclassifies CHAMPS cause $i$, the odds $\phi_{ij}/\phi_{ik}$ reflects its relative tendency to misclassify as causes $j$ versus $k$. If these odds depend only on $(j,k)$ and not on $i$, it indicates a systematic preference. This connects the base model to observable misclassification odds, enabling us to assess support for the base model for a given data.

#### S1.2 General Homogeneous Modeling of Misclassification Matrices

While the characterization from Theorem 3.1 in [1] provides evidence for the base model, the assumptions underlying the model may be restrictive for broader practical applications. The degree to which the base model assumptions hold could vary across age groups and VA algorithms. To address this, [1] proposes a model for a general unstructured misclassification matrix. It allows deviations from the base model and shrinks toward it for parsimony when either the sample size is limited or evidence is absent. Denote a general misclassification matrix as $\mathbf{\Phi} = (\phi_{ij})$ and the base model misclassification matrix as $\mathbf{\Phi}^{(b)} = (\phi_{ij}^{(b)})$ with $\phi_{ii}^{(b)} = a_i + (1-a_i)\rho_i$ and $\phi_{ij}^{(b)} = (1-a_i)\rho_j$. To characterize deviations from the base model, false negative rates are reparameterized via the exact decomposition $\phi_{ij} = (1 - \phi_{ii})q_{ij}$ for $i \neq j$ where $\phi_{ii}$ is the sensitivity and $q_{ij} = P(V = j|M = i,\ V \neq i)$ is the relative false negative, which satisfies $q_{ij} > 0$ and $\sum_{j\neq i} q_{ij} = 1$. Under the base model, the relative false negative simplifies to $q_{ij}^{(b)} = \rho_j/(1 - \rho_i)$.

The model independently places a Beta prior on the sensitivities $\phi_{ii}$ and a Dirichlet prior on the relative false negatives $q_{ij}$, where both are centered around their respective base model values $\phi_{ij}^{(b)}$ and $q_{ij}^{(b)}$. This approach assigns positive probability to every interior point within the space of $C \times C$ Markov matrices. Following notations in [1], let $\mathbf{T}_s = (t_{sij})$ be the observed misclassification matrix where $t_{sij}$ is the number of deaths classified as cause $j$ given true cause $i$ in country $s$, $\mathbf{T} = \sum_s \mathbf{T}_s$ is the pooled observed misclassification matrix, $\mathbf{T}_{si*}$, $\mathbf{T}_{i*}$, and $\mathbf{\Phi}_{i*}$ are $i^{th}$ row of $\mathbf{T}_s$, $\mathbf{T}$, and $\mathbf{\Phi}$, $\boldsymbol{q}_{i*} = (q_{i1}, \cdots, q_{i,i-1}, q_{i,i+1}, \cdots, q_{iC})^{\mathrm{T}}$, $\boldsymbol{q}_{i*}{}^{(b)} = (q_{i1}{}^{(b)}, \cdots, q_{i,i-1}{}^{(b)}, q_{i,i+1}{}^{(b)}, \cdots, q_{iC}{}^{(b)})^{\mathrm{T}}$, $\boldsymbol{\rho} = (\rho_1, \cdots, \rho_C)^{\mathrm{T}}$, $n_{si}$ is the number of cases in country $s$ with CHAMPS cause $i$, and $n_i = \sum_s n_{si}$ is the total sample size for CHAMPS cause $i$ across all countries. A general homogeneous model specifies

$$\mathbf{T}_{i*} \sim \text{Multinomial}(n_i, \mathbf{\Phi}_{i*}) \text{ independently with } \phi_{ij} = (1 - \phi_{ii})q_{ij} \text{ for } i \neq j,$$

$$\phi_{ii} \sim \text{Beta}\left(0.5 + \kappa\phi_{ii}^{(b)}, 0.5 + \kappa(1 - \phi_{ii}^{(b)})\right) \text{ independently with } \kappa > 0,$$

$$\boldsymbol{q}_{i*} \sim \text{Dirichlet}\left(0.5 + \lambda\boldsymbol{q}_{i*}{}^{(b)}\right) \text{ independently with } \lambda > 0,$$

$$a_{ii} \sim \text{Beta}(b, d) \text{ iid with } b, d > 0, \text{ and}$$

$$\boldsymbol{\rho} \sim \text{Dirichlet}(e_1, \cdots, e_C) \text{ with } e_j > 0.$$

With ample data, the posterior distribution concentrates around the true misclassification matrix. However, when data is scarce or sufficient evidence of deviation from the base model is absent, incorporating structured priors becomes essential to enhance estimation accuracy. As the hyperparameters $\kappa$ and $\lambda$ tend toward infinity, the model simplifies to the base model. Conversely, as $\kappa$ and $\lambda$ approach zero, the priors become Jeffreys' non-informative priors—specifically, $\text{Beta}(0.5{,}0.5)$ and $\text{Dirichlet}(0.5, \cdots, 0.5)$—which are often used in unstructured models for proportions. By tuning $\kappa$ and $\lambda$, the framework offers a flexible spectrum of models, spanning from highly structured misclassification under systematic preference to fully unstructured general misclassification formulations.

#### S1.3 County-Specific Modeling of Misclassification Matrices

As indicated by Figure 4 in the main article for neonates and Figure S25 below for children, observed CHAMPS-VA misclassification rates significantly vary across different countries. However, due to the limited availability of data per country, it is not feasible to naively estimate unstructured misclassification matrices for each country individually. The base model addresses this issue by capturing the underlying structure in the misclassification matrix, which significantly reduces dimensionality. This lets us incorporate cross-country heterogeneity in a Bayesian hierarchical framework, with country-specific estimates shrinking toward the general and baseline homogeneous misclassification matrix estimates.

Let $\mathbf{\Phi}_s = (\phi_{sij})$ denote the misclassification matrix for country $s$, and $\mathbf{T}_s$ is the observed misclassification matrix defined above. To capture country-level variation, we propose a heterogeneous model that places country-specific probabilistic models on sensitivities and relative false negatives, centered around the global (homogeneous) model as

$$\mathbf{T}_{si*} \sim \text{Multinomial}(n_{si}, \mathbf{\Phi}_{si*}) \text{ independently with } \phi_{sij} = (1 - \phi_{sii}) q_{sij} \text{ for } i \neq j,$$

$$\phi_{sii} \sim \text{Beta}(0.5 + \gamma\phi_{ii}, 0.5 + \gamma(1 - \phi_{ii})) \text{ independently with } \gamma > 0, \text{ and}$$

$$\boldsymbol{q}_{si*} \sim \text{Dirichlet}(0.5 + \delta \boldsymbol{q}_{i*}) \text{ independently with } \delta > 0.$$

The same priors as in Section S1.2 are specified for the pooled parameters $\phi_{ii}$ and $\boldsymbol{q}_{i*}$. To capture cross-country heterogeneity, $\phi_{sii}$ and $q_{sij}$ are treated as country-specific random effects and follow Beta and Dirichlet distributions, respectively. The distributions are centered around their corresponding fixed effects—i.e., the equivalent parameters $\phi_{ii}$ and $\boldsymbol{q}_{i*}$ in the homogeneous model. The deviation of random effects from the fixed effects reflects the extent of heterogeneity. The heterogeneous model generalizes the homogeneous model, reducing to it as $\gamma, \delta \uparrow \infty$. On the other hand, when $\gamma, \delta \downarrow 0$, the random effects independently follow Jeffreys' non-informative priors— $\mathrm{Beta}(0.5,0.5)$ and $\mathrm{Dirichlet}(0.5, \cdots, 0.5)$—which corresponds to modeling misclassification independently for each country without any information sharing. The framework thus also includes separate country-specific models as a limiting case. In intermediate cases for some $\gamma, \delta > 0$, where some heterogeneity is present, the fixed effects or homogeneous values represent average misclassification rates shared across countries, while the variability of the random effects captures country-specific deviations. The parameters $\gamma$ and $\delta$ thus allow the model to flexibly represent different degrees of heterogeneity within a unified hierarchical structure.

#### S1.4 Shrinkage Priors on Interpretable Effect Sizes for Parsimonious Modeling

The differences between the base homogeneous model, the general homogeneous model, and the country-specific model are regulated by the dispersion parameters $(\kappa, \lambda)$ and $(\gamma, \delta)$, respectively. They allow the framework to flexibly expand to non-informative priors or reduce to the parsimonious model from the previous hierarchy. In the Bayesian setting, when Beta and Dirichlet priors are used for Binomial and Multinomial likelihoods, the dispersion parameters can be interpreted as the total prior sample size. To reflect this interpretation, [1] parameterize the dispersion terms as:

$$\kappa = 2\omega_{\mathrm{P}}, \lambda = (C-1)\omega_{\mathrm{P}}, \gamma = 2\omega_{\mathrm{S}}, \text{ and } \delta = (C-1)\omega_{\mathrm{R}}.$$

Here $\omega_{\mathrm{P}}$, $\omega_{\mathrm{S}}$, and $\omega_{\mathrm{R}}$ are the effect sizes of the framework, and they are interpreted as prior sample sizes for each category. Together, $\omega_{\mathrm{P}}$ governs *pull strength* (shrinkage towards the base model), and $\omega_{\mathrm{S}}$ and $\omega_{\mathrm{R}}$ degrees of homogeneity for sensitivities and relative false negatives (shrinkage towards the general homogeneous model). Specifically, a larger $\omega_{\mathrm{P}}$ reflects a stronger pull, inducing greater shrinkage toward the base model that incorporates systematic preferences. Conversely, a smaller $\omega_{\mathrm{P}}$ value suggests minimal systematic preference and no shrinkage. Similarly, higher $\omega_{\mathrm{S}}$ indicates greater homogeneity by encouraging country-specific sensitivities $\phi_{sii}$ to align more closely with the homogeneous rates $\phi_{ii}$. $\omega_{\mathrm{R}}$ is similarly interpreted for relative false negatives $\boldsymbol{q}_{si*}$. Figure 4 in [1] provides a visual overview of the full country-specific misclassification modeling framework, illustrating how these effect sizes influence the model's behavior.

Given the critical role of the effect sizes in determining the pull strength and degree of homogeneity, [1] adopts a fully Bayesian perspective and specify shrinkage priors on effect sizes to promote parsimony. They specify $\mathrm{Beta}(\varepsilon, \varepsilon)$ prior on $f(\omega_{\mathrm{P}})$, $f(\omega_{\mathrm{S}})$, and $f(\omega_{\mathrm{R}})$ where $f(x) = 1/(1+x)$. This encourages the framework to favor simpler models when evidence is absent, yet allows a more complex behavior when supported by sufficient evidence. This allows the framework to dynamically adapt its complexity in a data-driven way, facilitating a balanced trade-off between bias and variance.

#### S1.5 Predicting Misclassification to Unobserved CHAMPS Causes and Countries

By leveraging information sharing across countries, the framework inherently supports the prediction of CHAMPS-VA misclassification rates for unobserved countries or CHAMPS causes. In the homogeneous model, the misclassification rates $\phi_{ii}$ for a new country follow the same posterior distribution as estimated from the training data, like from CHAMPS. The predictive distribution of misclassification rates $\phi_{sii}$ in a new country $s$ is given by

$$\phi_{sii}^{(r)} \sim \text{Beta}\left(0.5 + 2\omega_{\text{S}}^{(r)}\phi_{ii}^{(r)}, 0.5 + 2\omega_{\text{S}}^{(r)}(1 - \phi_{ii}^{(r)})\right) \text{ for all } i, \text{ and}$$

$$\phi_{sij}^{(r)} = \left(1 - \phi_{sij}^{(r)}\right)q_{sij}^{(r)} \text{ for } i \neq j \text{ where } \boldsymbol{q}_{si*}^{(r)} \sim \text{Dirichlet}\left(0.5 + (C-1)\omega_{\text{R}}^{(r)}\boldsymbol{q}_{i*}^{(r)}\right).$$

The predictive distributions are centered around the corresponding homogeneous estimates, with the associated uncertainty for sensitivity and relative false negatives informed by the posterior estimates of the effect sizes $\omega_{\text{S}}$ and $\omega_{\text{R}}$. The latter reflects the degree of homogeneity observed in the training. As a result, while the point estimates from the homogeneous and heterogeneous models tend to be similar, the heterogeneous model yields wider uncertainty intervals that reflect potential cross-country heterogeneity.

#### S1.6 Modular VA-Calibration Using Uncertainty-Quantified Country-Specific Misclassification Estimates

A key objective in refining CHAMPS-VA misclassification estimation is to improve the accuracy of Cause-Specific Mortality Fraction (CSMF) estimates when calibrating VA-only cause of death (COD) data from a target country, such as COMSA-Mz in Mozambique. For this, [1] proposed an approximate modularized VA-Calibration framework. Let $\mathcal{U} \coloneqq \mathcal{U}_s$ represent the

VA-only single-cause COD data $\{\boldsymbol{V}_r\}_{r\in\mathcal{U}}$ of $N$ deaths from a target country $u$ and $\boldsymbol{p} = (p_1, \cdots, p_C)^{\mathrm{T}}$ denote the CSMF in the country. Here $\boldsymbol{V}_r = (V_{r1}, \cdots, V_{rC})^{\mathrm{T}}$ , with $V_{rj} = 1$ if and only if COD is cause $j$ and $\sum_{j=1}^{C} V_{rj} = 1$ for all $r$. The uncalibrated CSMF estimate equals to $\widehat{\boldsymbol{p}} = (\hat{p}_1, \cdots, \hat{p}_C)^{\mathrm{T}}$ where $\hat{p}_j = \sum_{r=1}^{N} V_{rj}/N$, which is simply the average (equal weights) of the uncalibrated estimates produced by the individual algorithms. Additionally, let $\mathcal{L}$ denote the labeled (paired) CHAMPS-VA COD data across countries. First, the approach analyzes the labeled dataset $\mathcal{L}$ once using the framework in Section S1.3, and stores posterior and predictive samples of country-specific misclassification matrices. For calibrating VA-only COD data from any country $s$, the modularized VA-Calibration specifies the following Bayesian hierarchical model:

$$\boldsymbol{V}_r \sim \text{Multinomial}\left(1, \boldsymbol{\Phi}_{si}^{\mathrm{T}}\boldsymbol{p}\right) \text{ iid for all } r \in \mathcal{U},$$

$$\boldsymbol{\Phi}_{si*} \sim \text{Dirichlet}(\boldsymbol{e}_{si}) \text{ independently for all } i,\ \boldsymbol{e}_{si} = (e_{si1}, \cdots, e_{siC})^{\mathrm{T}} \text{ and } e_{sij} > 0,$$

$$\boldsymbol{p} \sim \text{Dirichlet}(1 + C\eta\widehat{\boldsymbol{p}}) \text{ with } \eta > 0.$$

Here, $\boldsymbol{e}_{si}$ represents the parameters of the Dirichlet distribution that best approximates the marginal posterior (if the country $s$ is observed in the labeled data) or predictive distribution (if the country $s$ is not observed in the labeled data) of $\boldsymbol{\Phi}_{si*}$ (misclassification of the algorithm in the country $s$ given CHAMPS cause $i$). The prior on $\boldsymbol{p}$ is the same as the current VA-calibration implementation, which introduces shrinkage toward the uncalibrated CSMF estimate $\widehat{\boldsymbol{p}}$ under limited samples. The degree of shrinkage is governed by $\eta$, which functions analogously to the effect sizes $\omega_{\mathrm{P}}$, $\omega_{\mathrm{S}}$, and $\omega_{\mathrm{R}}$. Following [1], we fix $\eta = 4$ for the analysis presented here.

For ensemble calibration, the above approach is extended to combine multiple CCVA algorithms. The modularized ensemble VA-Calibration for $K$ CCVA algorithms specifies:

$$\boldsymbol{V}_{kr} \sim \text{Multinomial}\left(1, \boldsymbol{\Phi}_{ksi}^{\mathrm{T}}\boldsymbol{p}\right) \text{ iid for all } r \in \mathcal{U} \text{ and } k = 1, \cdots, K,$$

$$\boldsymbol{\Phi}_{ksi*} \sim \text{Dirichlet}(\boldsymbol{e}_{ksi}) \text{ independently for all } i, \boldsymbol{e}_{ksi} = (e_{ksi1}, \cdots, e_{ksiC})^{\mathrm{T}} \text{ and } e_{ksij} > 0,$$

$$\boldsymbol{p} \sim \text{Dirichlet}(1 + C\eta\widehat{\boldsymbol{p}}) \text{ with } \eta > 0.$$

Here, $\mathcal{U}_k$ represent the VA-only single-cause COD data $\{\boldsymbol{V}_{kr}\}_{r\in\mathcal{U}_k}$ of $N_k$ deaths, $V_{krj}$ is the predicted COD for an individual $r$ using algorithm $k$ with $V_{krj} = 1$ if and only if COD is cause $j$ and $\sum_{j=1}^{C} V_{krj} = 1$ for all $k$ and $r$. $\boldsymbol{e}_{ksi}$ represents the parameters of the Dirichlet distribution that best approximates the marginal posterior (if country $s$ is observed in the labeled data) or predictive distribution (if the country $s$ is not observed in the labeled data) of $\boldsymbol{\Phi}_{ksi*}$ (misclassification of algorithm $k$ in country $s$ given CHAMPS cause $i$) obtained from the analysis of CHAMPS data. $\widehat{\boldsymbol{p}} = (\hat{p}_1, \cdots, \hat{p}_C)^{\mathrm{T}}$ here is the ensemble uncalibrated CSMF estimate where $\hat{p}_j = \sum_{k=1}^{K}\sum_{r=1}^{N_k} V_{krj}/\sum_{k=1}^{K} N_k$.

In many situations, we may not want to calibrate all causes. For example, 'other' cause is not calibrated in analyzing Mozambique's CSMF because 'other' cause in CHAMPS and CCVA algorithms consists of different collections of finer ICD 10-level causes. The above calibration framework can be adjusted accordingly for this. Without loss of generality, consider one algorithm and suppose that cause 1 need not be calibrated. Note that, we can write $\phi_{sij} = (1 - \phi_{si1})\phi_{sij}^{\dagger}$ for $i, j = 2, \cdots, C$, $p_j = (1 - p_1)p_j^{\dagger}$, and $\hat{p}_j = (1 - \hat{p}_1)\hat{p}_j^{\dagger}$ for $j = 2, \cdots, C$, where $\phi_{sij}^{\dagger}$, $p_j^{\dagger}$, $\hat{p}_j^{\dagger}$ are renormalized $\phi_{sij}$, $p_j$, $\hat{p}_j$ after removing cause 1 and $\boldsymbol{e}_{si}^{\dagger} = (e_{si2}, \cdots, e_{siC})^{\mathrm{T}}$. Let us also denote by $\mathcal{U}^{\dagger}$ obtained from $\mathcal{U}$ after removing deaths from cause 1. Following the renormalizing rule of the Dirichlet distribution we have $\boldsymbol{\Phi}_{si*}^{\dagger} = (\phi_{si2}^{\dagger}, \cdots, \phi_{siC}^{\dagger})^{\mathrm{T}} \sim \text{Dirichlet}\left(\boldsymbol{e}_{si}^{\dagger}\right)$

independently for all $i = 2, \cdots, C$. Denote $\boldsymbol{p}^\dagger = (p_2^\dagger, \cdots, p_C^\dagger)^\mathrm{T}$ and $\widehat{\boldsymbol{p}}^\dagger = (\hat{p}_2^\dagger, \cdots, \hat{p}_C^\dagger)^\mathrm{T}$. To calibrate for causes 2 to $C$, we specify the following Bayesian hierarchical model:

$$\boldsymbol{V}_r \sim \text{Multinomial}\left(1, (\boldsymbol{\Phi}_{si}^\dagger)^\mathrm{T} \boldsymbol{p}^\dagger\right) \text{ iid for all } r \in \mathcal{U}^\dagger,$$

$$\boldsymbol{\Phi}_{si*}^\dagger \sim \text{Dirichlet}\left(\boldsymbol{e}_{si}^\dagger\right) \text{ independently for all } i,$$

$$\boldsymbol{p}^\dagger \sim \text{Dirichlet}(1 + C\eta \widehat{\boldsymbol{p}}^\dagger) \text{ with } \eta > 0.$$

Here $\boldsymbol{\Phi}_{si*}^\dagger$ are rows of $\boldsymbol{\Phi}_{si}^\dagger$. Then $r^{th}$ posterior sample of calibrated CSMF $p_j^{(r)}$ is given by $p_1^{(r)} = \hat{p}_1$ and $p_j^{(r)} = (1 - \hat{p}_1) {p^\dagger}_j^{(r)}$ for $j = 2, \cdots, C$, where ${p^\dagger}_j^{(r)}$ is the $r^{th}$ posterior sample of calibrated CSMF $p_j^\dagger$. This can be similarly extended to ensemble calibration.

Here, we presented a brief overview of the country-specific misclassification matrix modeling framework, and the modular VA-Calibration approach proposed in [1], which were employed for the analysis presented here. For further details, please refer to the original publication and its accompanying supplementary materials.

#### S2. Neonates (0-27 days)

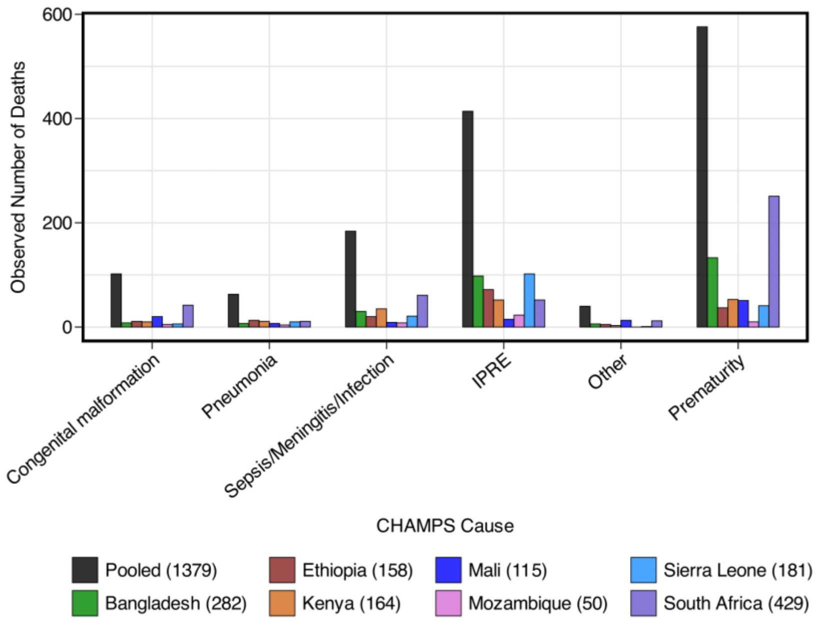


Figure S1. Observed number of deaths for different CHAMPS causes for neonates (0-27 days). "Pooled" (black) on the far left combines deaths from all countries. Parentheses in the legend indicate the number of deaths observed in each country. IPRE denotes intrapartum-related events.

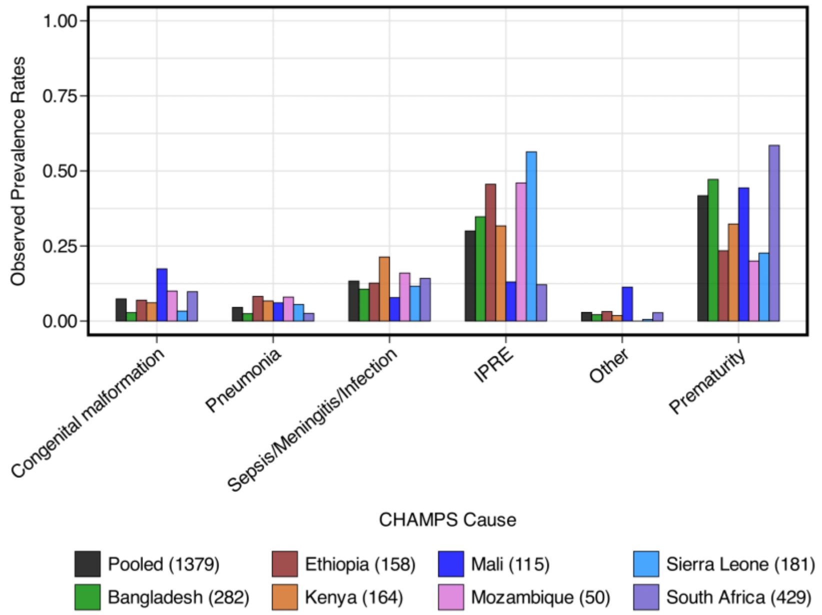


Figure S2. Observed proportion of deaths for different CHAMPS causes for neonates (0-27 days). "Pooled" (black) on the far left combines deaths from all countries. Parentheses in the legend indicate the number of deaths observed in each country. IPRE denotes intrapartum-related events.

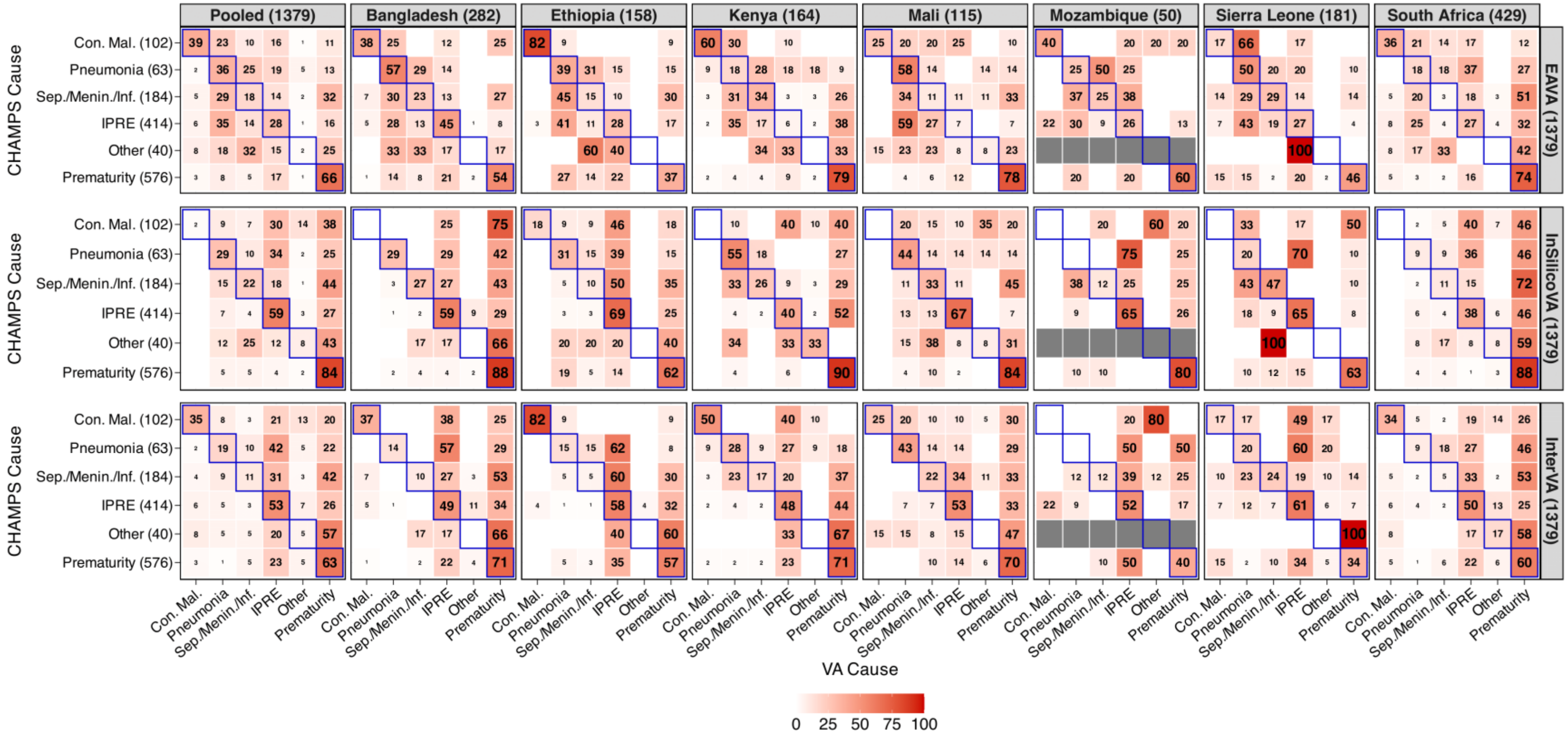

Figure S3. **Observed** misclassification rates in % of EAVA (top row), InSilicoVA (middle row), and InterVA (bottom row) for neonatal deaths (0-27 days) in CHAMPS. Pooled rates in the far-left column are calculated by combining rates across all countries. Parentheses in the column headers at the top indicate the number of deaths per country. Parentheses in the row names on the left represent the total death counts for each CHAMPS cause combined across countries. Parentheses in the panel names on the right show the total death counts for each CCVA algorithm. Sensitivities are along diagonals (outlined in blue) for each country. Grey cells indicate the CHAMPS cause was not observed in that country. Con. mal., Sep./Menin./Inf., IPRE denote congenital malformation, sepsis/meningitis/infection, and intrapartum-related events.

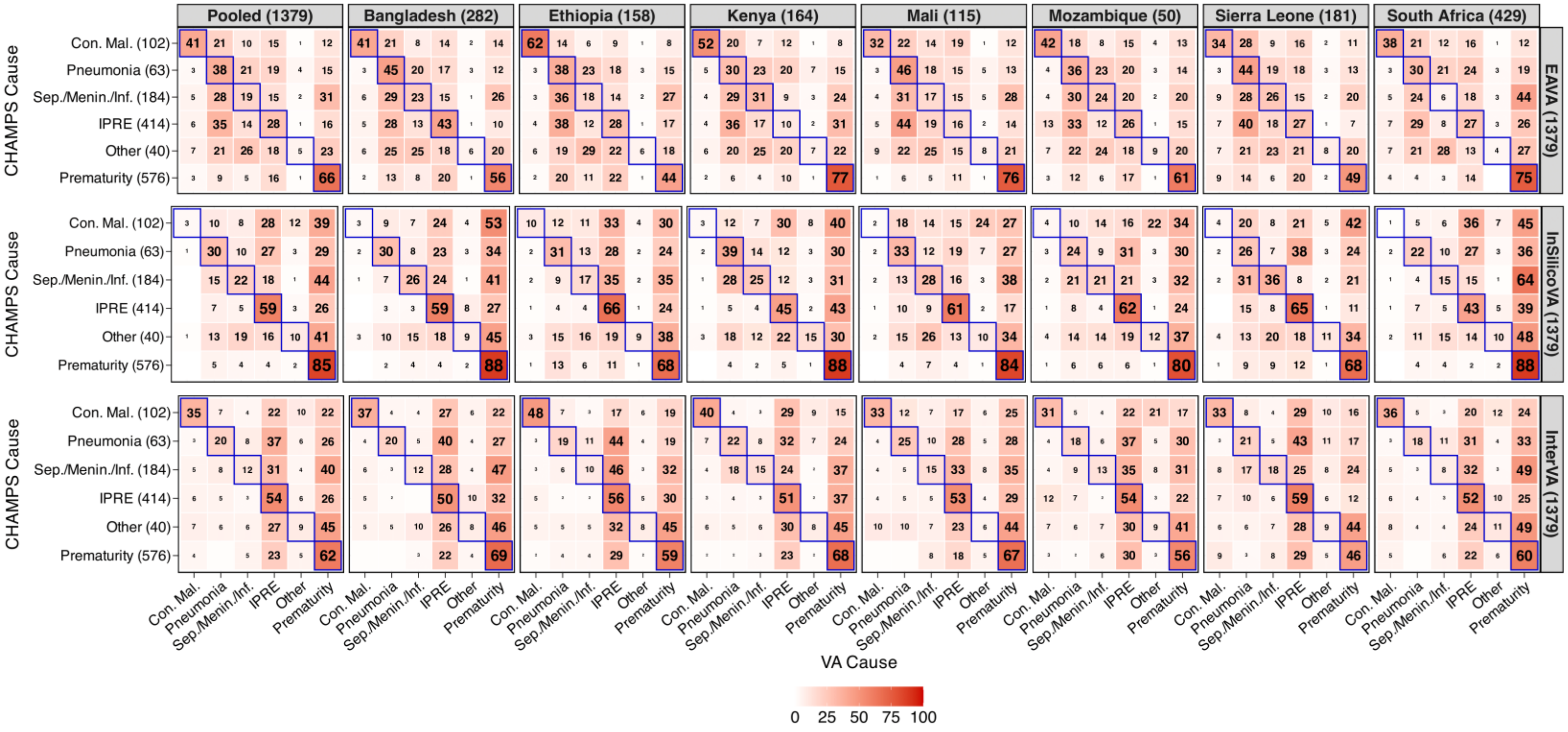

Figure S4. **Estimated** misclassification rates in % of EAVA (top row), InSilicoVA (middle row), and InterVA (bottom row) for neonatal deaths (0-27 days) in CHAMPS. Pooled rates in the far-left column are rates combined across all countries. Parentheses in the column headers at the top indicate the number of deaths per country. Parentheses in the row names on the left represent the total death counts for each CHAMPS cause combined across countries. Parentheses in the panel names on the right show the total death counts for each CCVA algorithm. Sensitivities are along diagonals (outlined in blue) for each country. Grey cells indicate the CHAMPS cause was not observed in that country. Con. mal., Sep./Menin./Inf., IPRE denote congenital malformation, sepsis/meningitis/infection, and intrapartum-related events.

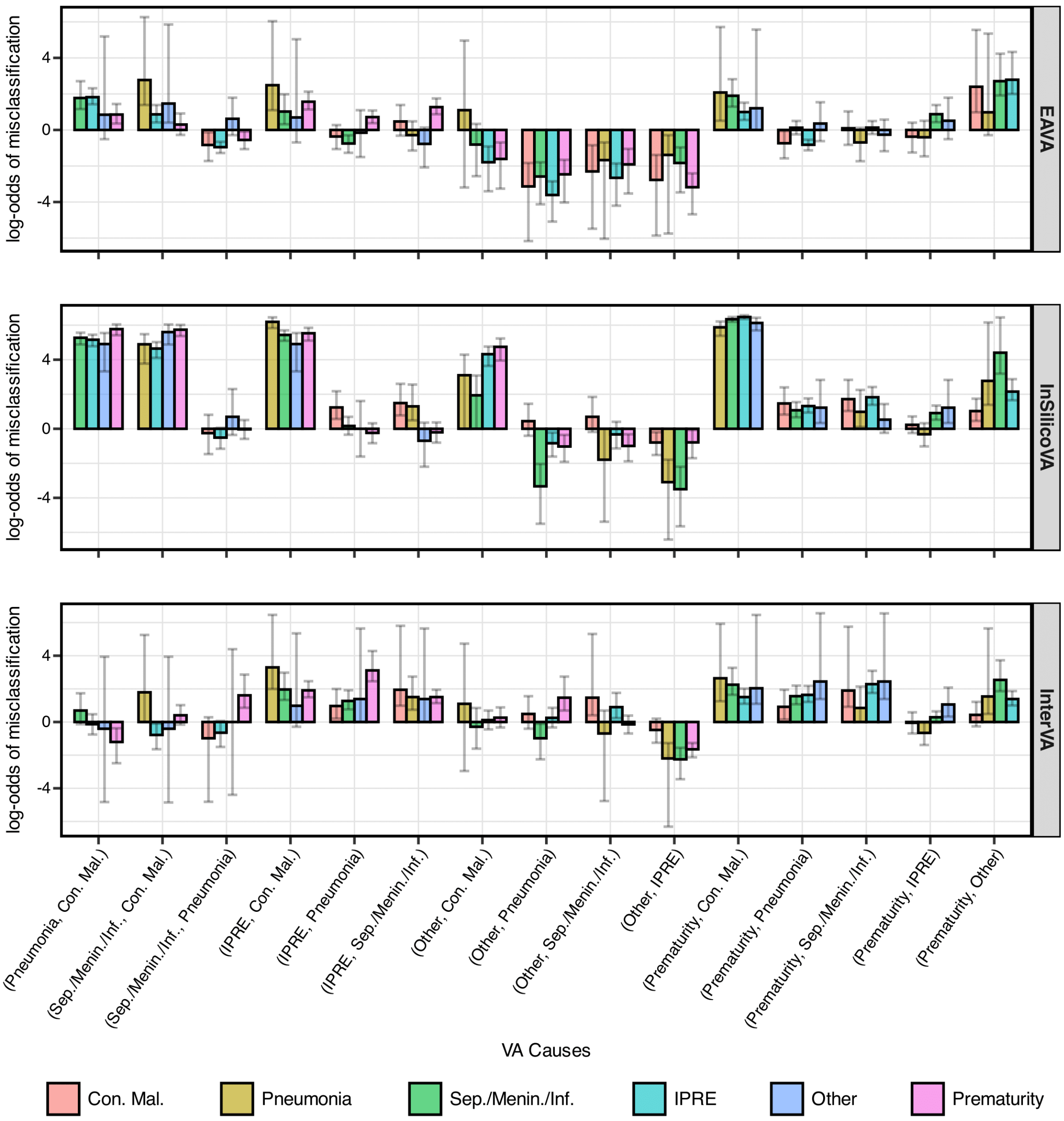


Figure S5. Grouped bar plot showing the similarity of log misclassification odds between CHAMPS causes for neonatal deaths (0-27 days). For each VA cause pair on the horizontal axis, log-odds values for corresponding CHAMPS causes are represented by color-coded bars on the vertical axis. Error bars indicate 95% confidence intervals calculated from 100,000 bootstrap samples. Con. mal., Sep./Menin./Inf., IPRE denote congenital malformation, sepsis/meningitis/infection, and intrapartum-related events. The heights of the bars within each group are roughly similar. This suggests strong evidence favoring the base model proposed in [1]. This is following the log-odds characterization according to Theorem 3.1 from [1].

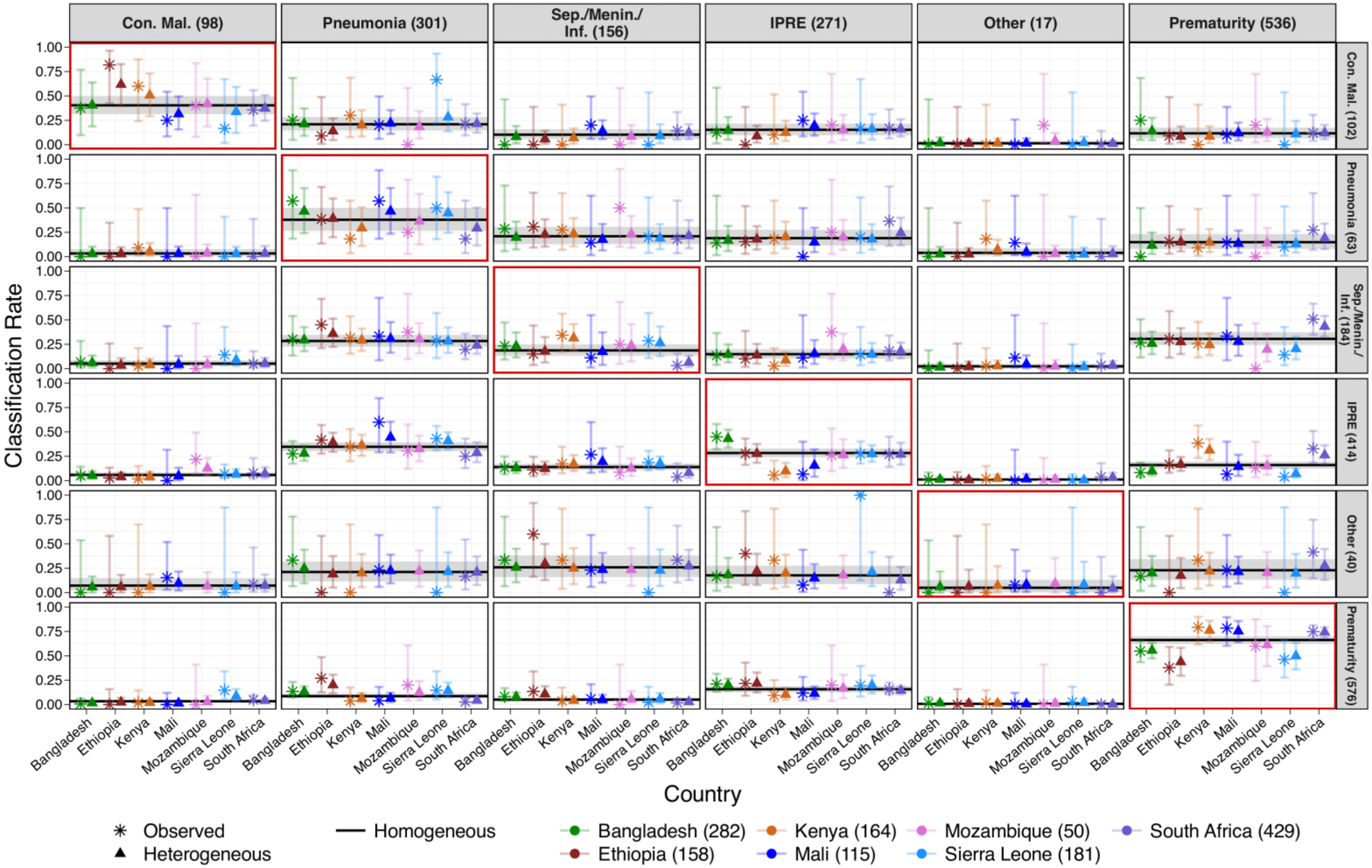


Figure S6. Comparison of observed **EAVA** misclassification rates with in-sample point (posterior mean) and uncertainty (95% credible intervals) estimates from homogeneous and country-specific models for neonates (0-27 days) in CHAMPS. Rows and columns are to CHAMPS and VA causes, with combined sample sizes across countries indicated in parentheses. Misclassification rates are conditioned on CHAMPS cause (row), so values in each row sum to 1 for each country and method. Con. mal., Sep./Menin./Inf., IPRE denote congenital malformation, sepsis/meningitis/infection, and intrapartum-related events.

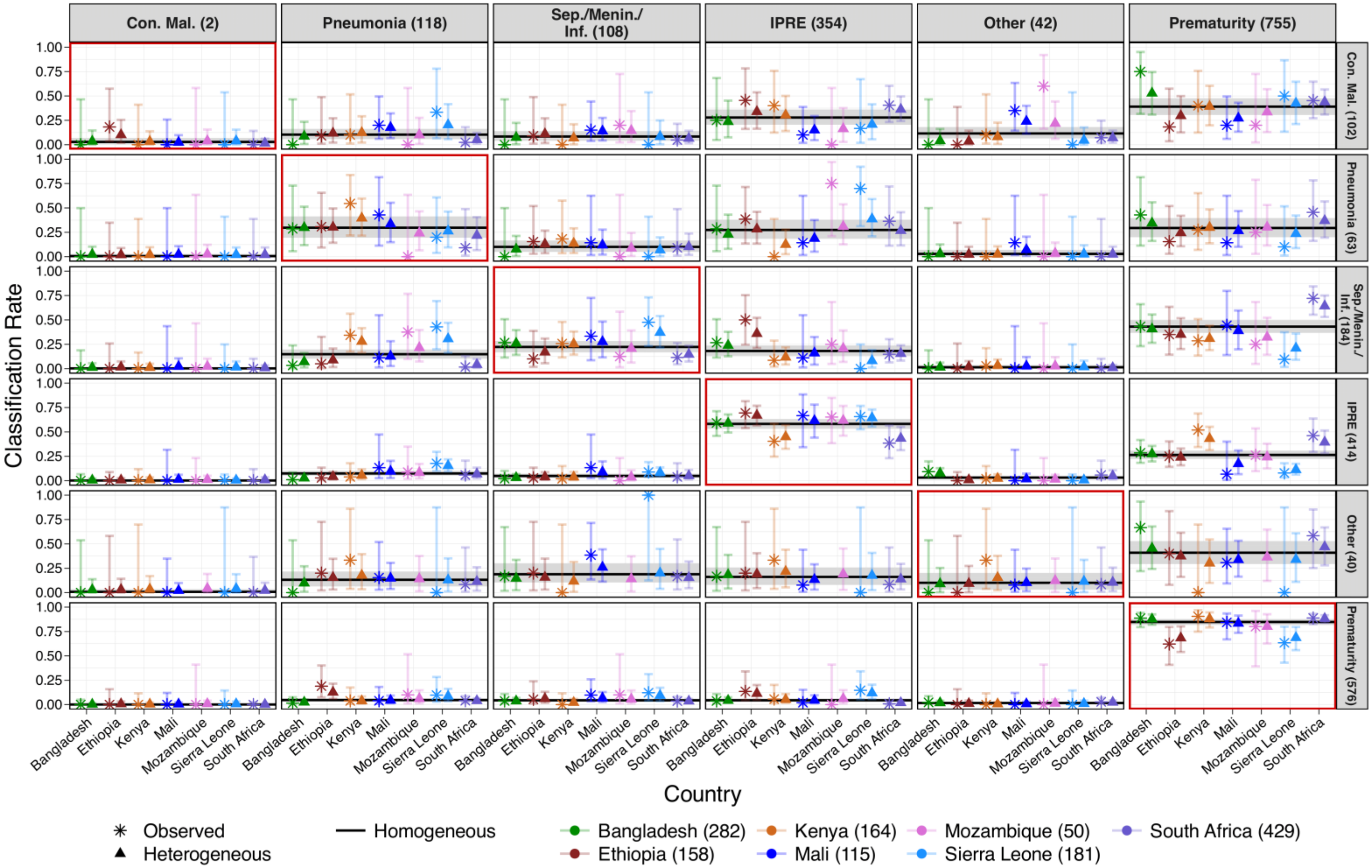


Figure S7. Comparison of observed **InSilicoVA** misclassification rates with in-sample point (posterior mean) and uncertainty (95% credible intervals) estimates from homogeneous and country-specific models for neonates (0-27 days) in CHAMPS. Rows and columns are to CHAMPS and VA causes, with combined sample sizes across countries indicated in parentheses. Misclassification rates are conditioned on CHAMPS cause (row), so values in each row sum to 1 for each country and method. Con. mal., Sep./Menin./Inf., IPRE denote congenital malformation, sepsis/meningitis/infection, and intrapartum-related event.

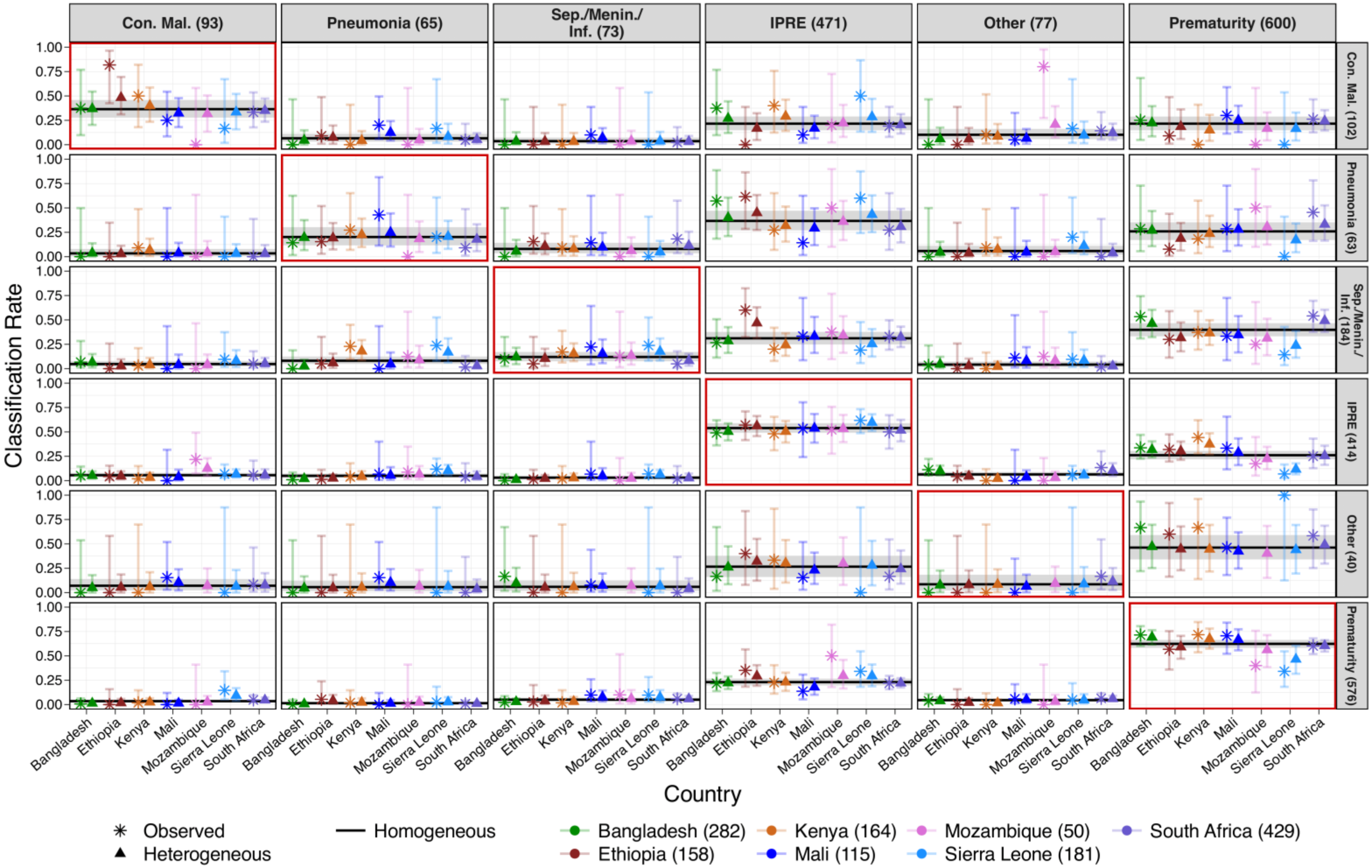


Figure S8. Comparison of observed **InterVA** misclassification rates with in-sample point (posterior mean) and uncertainty (95% credible intervals) estimates from homogeneous and country-specific models for neonates (0-27 days) in CHAMPS. Rows and columns are to CHAMPS and VA causes, with combined sample sizes across countries indicated in parentheses. Misclassification rates are conditioned on CHAMPS cause (row), so values in each row sum to 1 for each country and method. Con. mal., Sep./Menin./Inf., IPRE denote congenital malformation, sepsis/meningitis/infection, and intrapartum-related event.

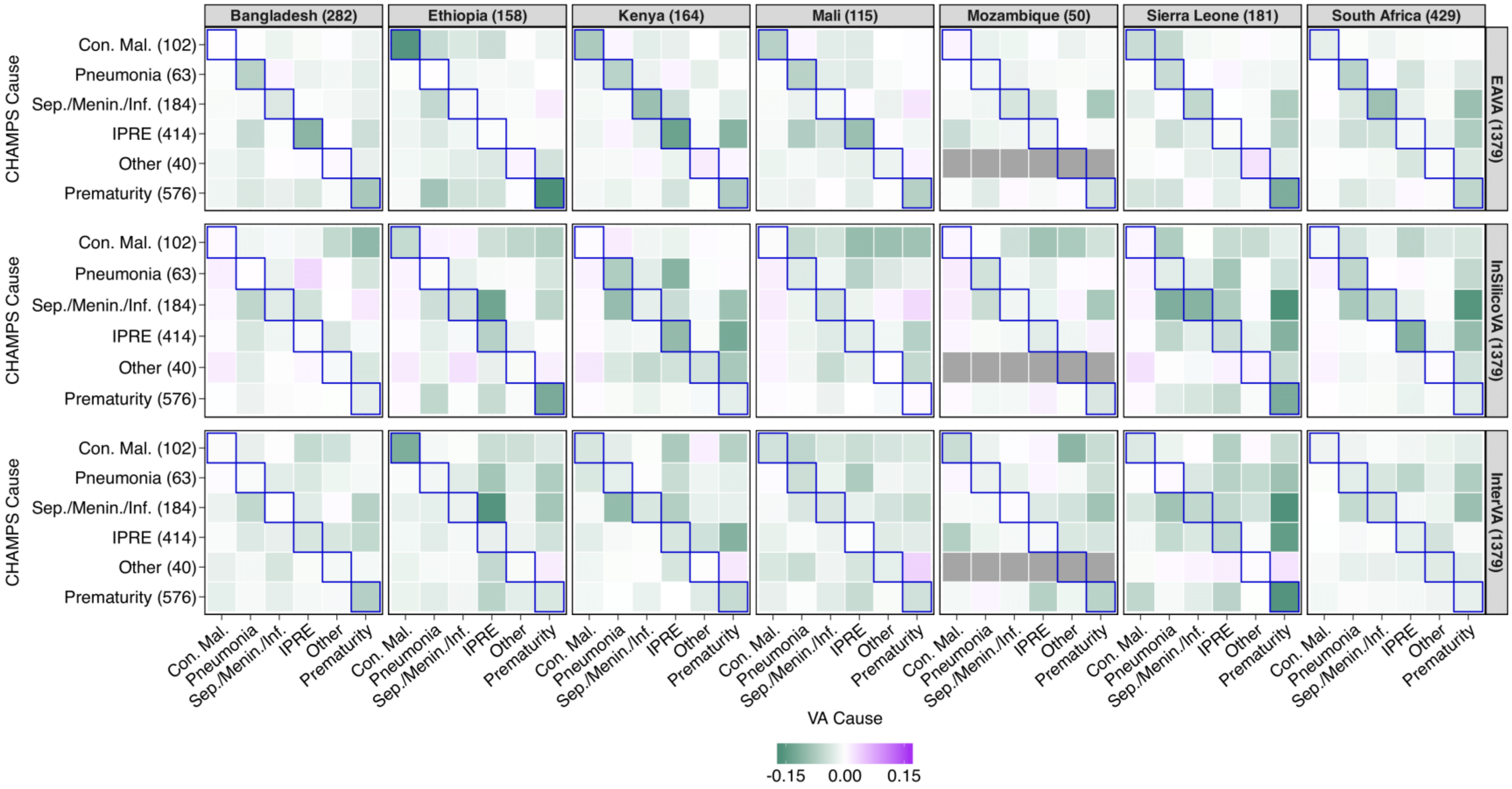


Figure S9. The change in the absolute in-sample bias of point estimates in the country-specific model compared to the homogeneous model among neonates (0-27 days). Rows and columns are CHAMPS and VA causes. Negative values (green color) are better and indicate a reduction in absolute in-sample bias. Grey cells indicate the CHAMPS cause was not observed for that country. Con. mal., Sep./Menin./Inf., IPRE denote congenital malformation, sepsis/meningitis/infection, and intrapartum-related events. Combined across countries, the country-specific model reduces bias for 83%, 72%, and 91% cause-pairs for EAVA, InSilicoVA, and InterVA with 2-22%, 3-22%, 3-16% improvements (lower bias) for half of them.

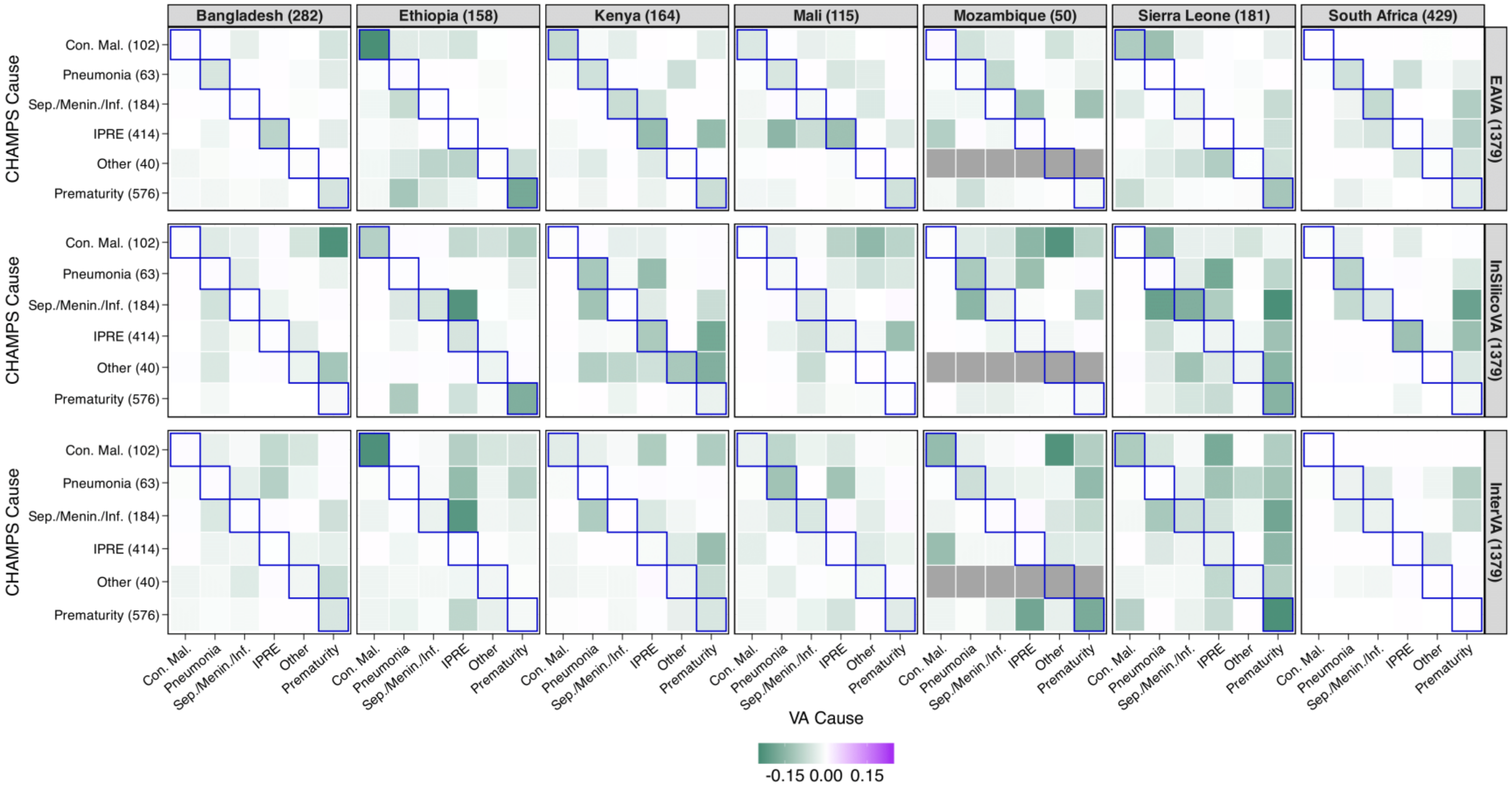


Figure S10. The change in interval scores of 95% credible intervals in the country-specific model compared to the homogeneous model among neonates (0-27 days). Rows and columns are CHAMPS and VA causes. Negative values (green color) are better and indicate a reduction in absolute interval scores. Grey cells indicate the CHAMPS cause was not observed for that country. Con. mal., Sep./Menin./Inf., IPRE denote congenital malformation, sepsis/meningitis/infection, and intrapartum-related events. Combined across countries, the country-specific model reduces bias for 83%, 72%, and 91% cause-pairs for EAVA, InSilicoVA, and InterVA with 3-31%, 4-27%, 2-23% improvements (lower scores) for half of them.

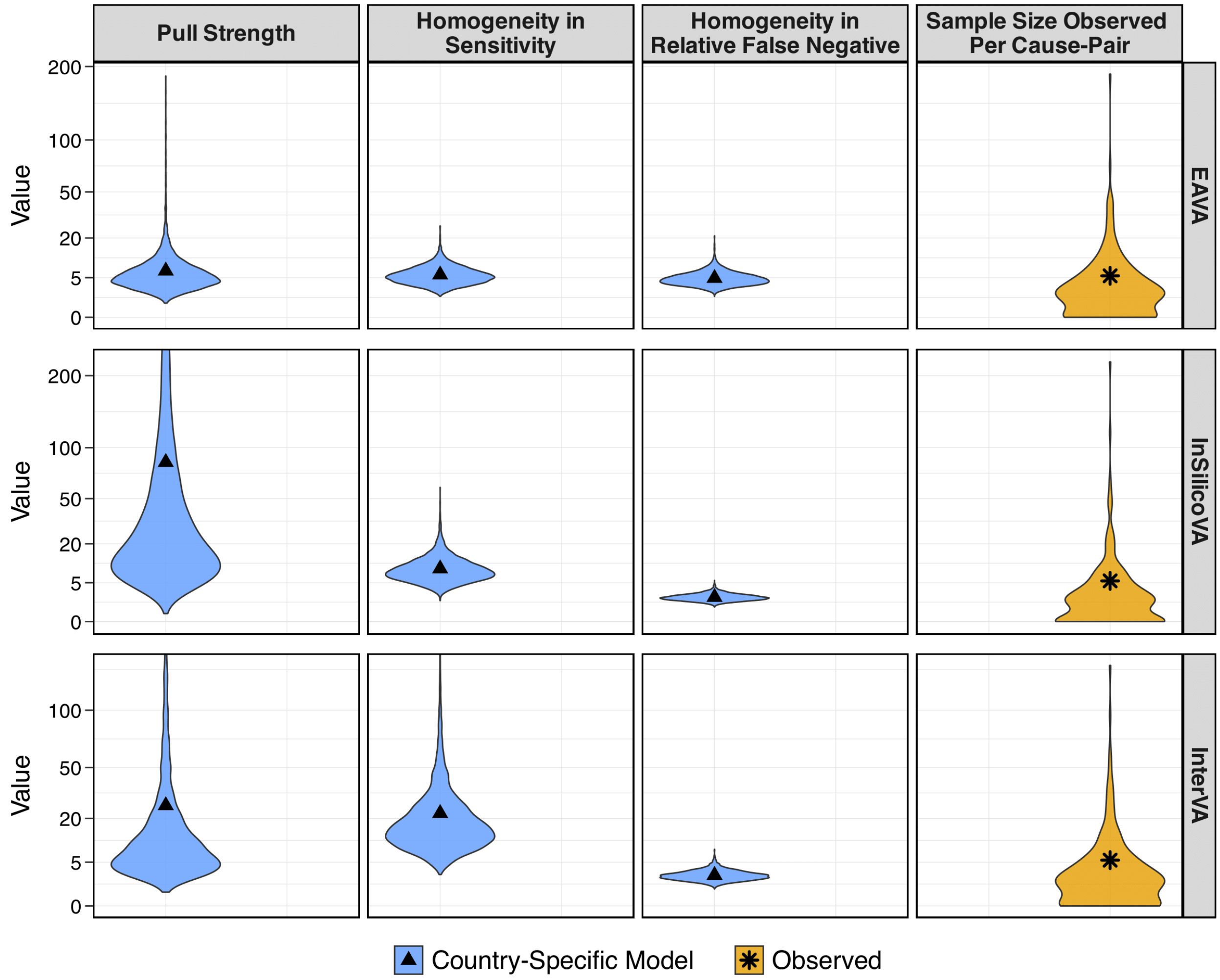


Figure S11. The first three columns present uncertainty-quantified estimates of effect sizes $\omega_P$ (pull strength), $\omega_S$ (degrees of heterogeneity in sensitivity), and $\omega_R$ (degrees of heterogeneity in relative false negative) in VA misclassification among neonatal (0-27 days) deaths in CHAMPS. The rightmost column shows the distribution of sample sizes across cause pairs and countries to compare effect size magnitude. Higher values relative to the observed sample size indicate stronger effects. Black points represent the means of each distribution. For clarity, the y-axis is scaled on the square root scale.

Figure S11 illustrates the effect size estimates (posterior densities; black triangles indicate posterior mean) of $\omega_{\mathrm{P}}$ (pull strength), $\omega_{\mathrm{S}}$ (degree of homogeneity in sensitivity), and $\omega_{\mathrm{R}}$ (degree of homogeneity in relative false negative) from the country-specific model. Since effect sizes are interpreted as prior sample sizes for each cause or category, their magnitude must be considered alongside the observed sample sizes across cause pairs and countries (last column; black stars indicate average). First, the estimates (first column) of pull strength for EAVA, InSilicoVA, and InterVA are 7, 84, and 26, respectively, compared to an average observed sample size of 5. This indicates that the strongest evidence for the base model is found in InSilicoVA, followed by InterVA, with EAVA showing the weakest support. Second, the estimates (second column) for the degree of homogeneity in sensitivity are 6 for EAVA, 9 for InSilicoVA, and 22 for InterVA. Relative to the average observed sample size of 5, this suggests that InterVA exhibits the most homogeneous sensitivities, followed by InSilicoVA, while EAVA displays the greatest heterogeneity in sensitivity. Lastly, the estimates (third column) for the degree of homogeneity in relative false negatives are 5, 2, and 2 for the three algorithms. Compared to the average observed sample size, this suggests considerable heterogeneity in relative false negativity compared to sensitivity, with the least heterogeneity for EAVA and the highest heterogeneity for InSilicoVA and InterVA. These findings point to varying degrees of systematic behavior and heterogeneity across the different CCVA algorithms and demonstrate the misclassification model's ability to capture and account for these differences adaptively in providing more accurate misclassification matrix estimates.

### S3. Children (1-59 months)

#### S3.1 Estimates of VA Misclassification Rates in CHAMPS

**VA Misclassification rates.** Misclassification rates for the CCVA algorithms were estimated using data from 1,080 recorded child deaths in CHAMPS, as shown in Figure S14. Using this, the method from [1] provides country-specific misclassification rates for each algorithm, along with quantified uncertainties. For simplicity, only point estimates are presented in Figure S15; full estimates with uncertainty intervals are in Figure S16-Figure S18.

Based on pooled rates in Figure S14 and Figure S15, we find significant variability in sensitivities across causes, even when aggregated across countries. According to the modeled rates in Figure S15, sensitivity is highest for injury (74-82%) and lowest for severe malnutrition (6-13%) and other (0-14%). Sensitivity also differs across algorithms. For example, InSilicoVA and InterVA have very low sensitivity for neonatal causes (2-3%), whereas EAVA's sensitivity reaches 37%. Similarly, for pneumonia, EAVA and InSilicoVA have sensitivities of around 41-46%, compared to about 23% for InterVA. Pooled false negative rates are also notable; for instance, for CHAMPS cause malaria, the false negative rate when the VA cause is classified as other infection is around 25-30% across algorithms. False negative rates vary by algorithm; for example, for CHAMPS causes severe malnutrition, EAVA and InSilicoVA have about 24-25% false negative rates for the VA cause pneumonia, while InterVA's rate is much lower, at around 9%.

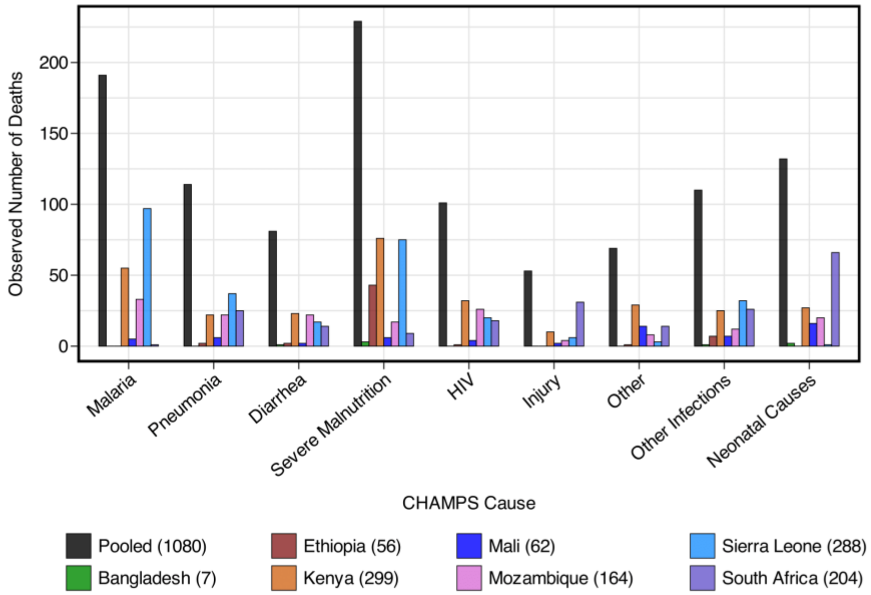


Figure S12. Observed number of deaths for different CHAMPS causes rates for children (1-59 months). "Pooled" (black) on the far-left combines deaths from all countries. Parentheses in the legend indicate the number of deaths observed in each country.

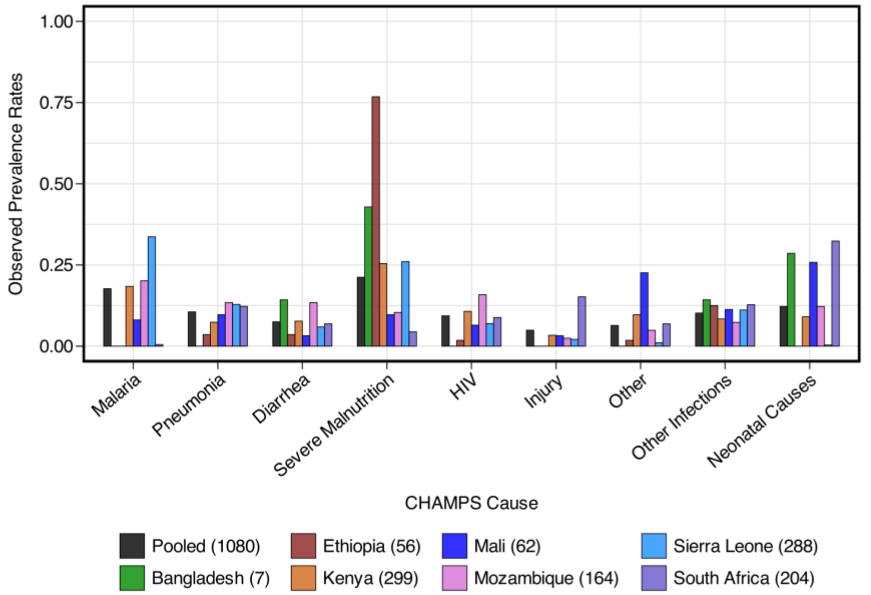


Figure S13. Observed proportion of deaths for different CHAMPS causes rates for children (1-59 months). "Pooled" (black) on the far-left combines deaths from all countries. Parentheses in the legend indicate the number of deaths observed in each country.

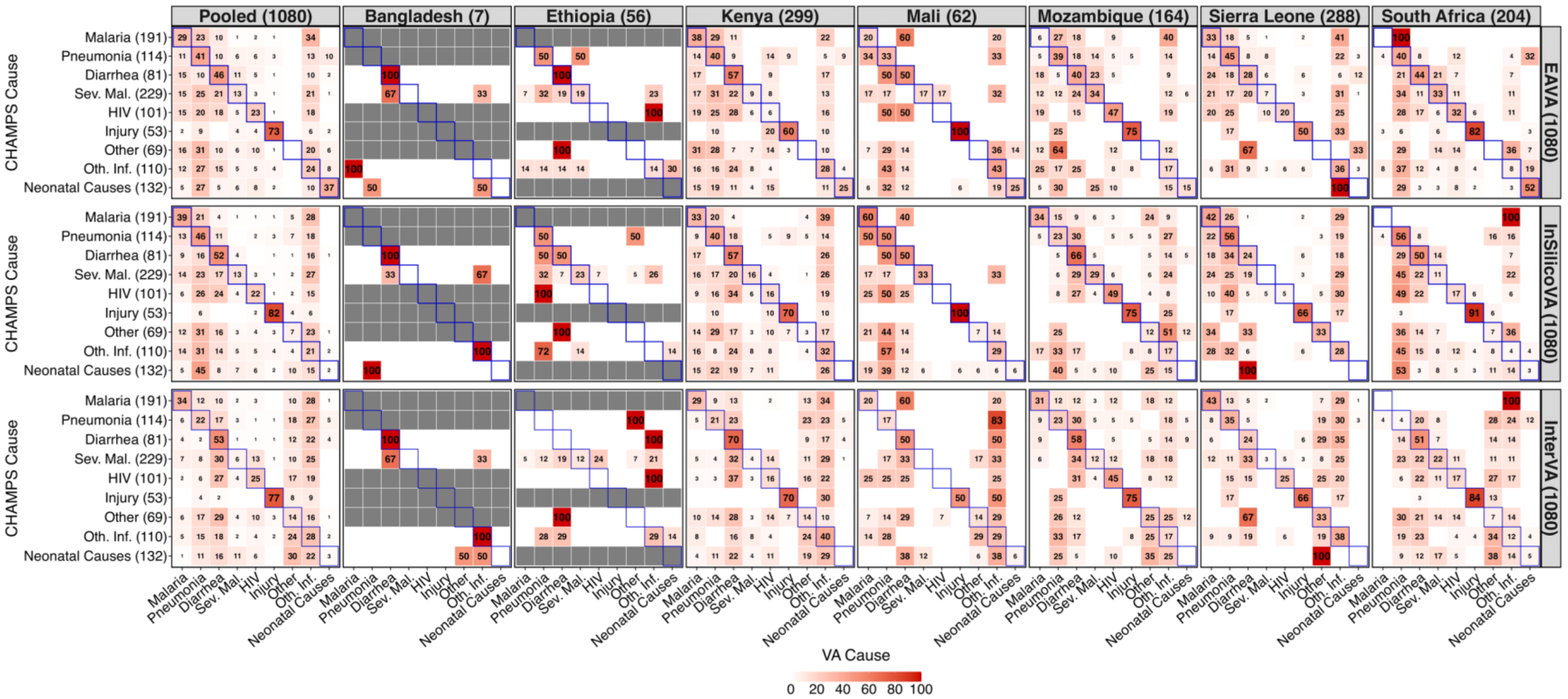

Figure S14. Observed misclassification rates in % of EAVA (top row), InSilicoVA (middle row), and InterVA (bottom row) for child deaths (1-59 months) in CHAMPS data. Pooled rates on the far-left column are calculated by combining rates across all countries. Parentheses in the column headers at the top indicate the number of deaths per country. Parentheses in the row names on the left represent the total death counts for each CHAMPS cause combined across countries. Parentheses in the panel names on the right show the total death counts for each CCVA algorithm. Sensitivities are along diagonals (outlined in blue) for each country. Grey cells indicate the CHAMPS cause was not observed for that country. Sev. mal. and Oth. Inf. denote severe malnutrition and other infections.

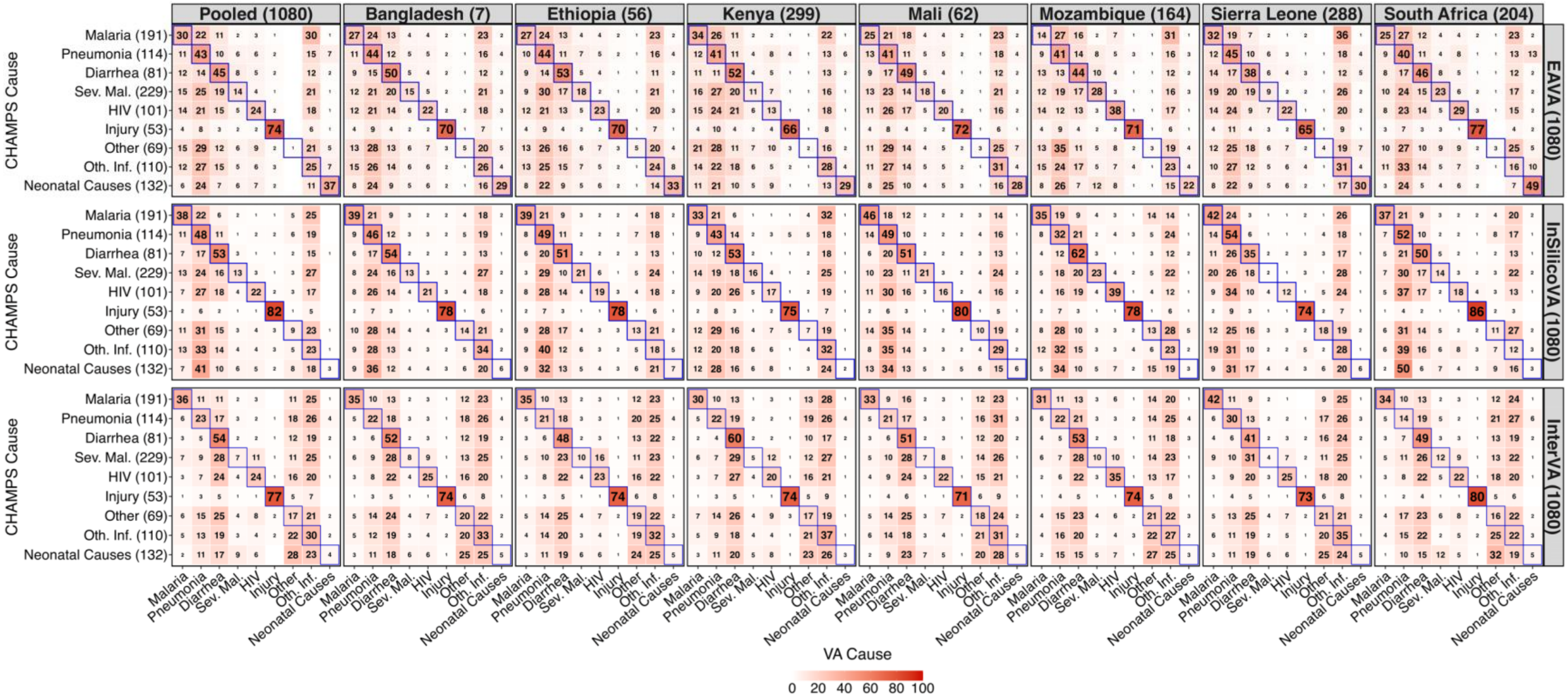


Figure S15. Estimated misclassification rates in % of EAVA (top row), InSilicoVA (middle row), and InterVA (bottom row) for child deaths (1-59 months) in CHAMPS data. Pooled rates in the far-left column are rates combined across all countries. Parentheses in the column headers at the top indicate the number of deaths per country. Parentheses in the row names on the left represent the total death counts for each CHAMPS cause combined across countries. Parentheses in the panel names on the right show the total death counts for each CCVA algorithm. Sensitivities are along diagonals (outlined in blue) for each country. Sev. mal. and Oth. Inf. denote severe malnutrition and other infections.

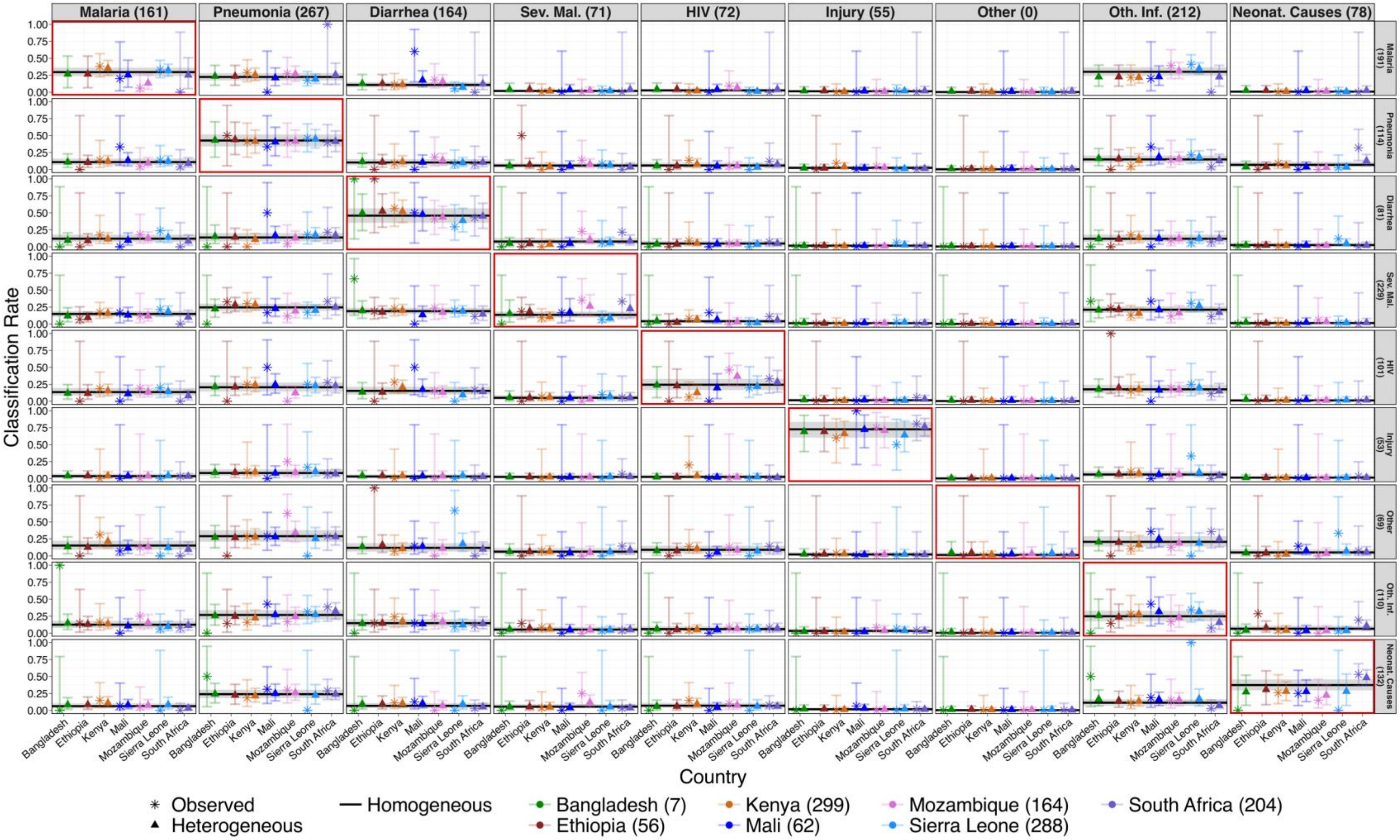


Figure S16. Comparison of observed misclassification rates with in-sample point (posterior mean) and uncertainty (95% credible intervals) estimates from homogeneous and heterogeneous models for EAVA among child (1-59 months) deaths in CHAMPS. Rows and columns correspond to CHAMPS and VA causes, with combined sample sizes across countries indicated in parentheses. Misclassification rates are conditioned on CHAMPS cause (row), so values in each row sum to 1 for each country and method. Sev. mal., Oth. Inf., and Neonat. Causes denote severe malnutrition, other infections, and neonatal causes.

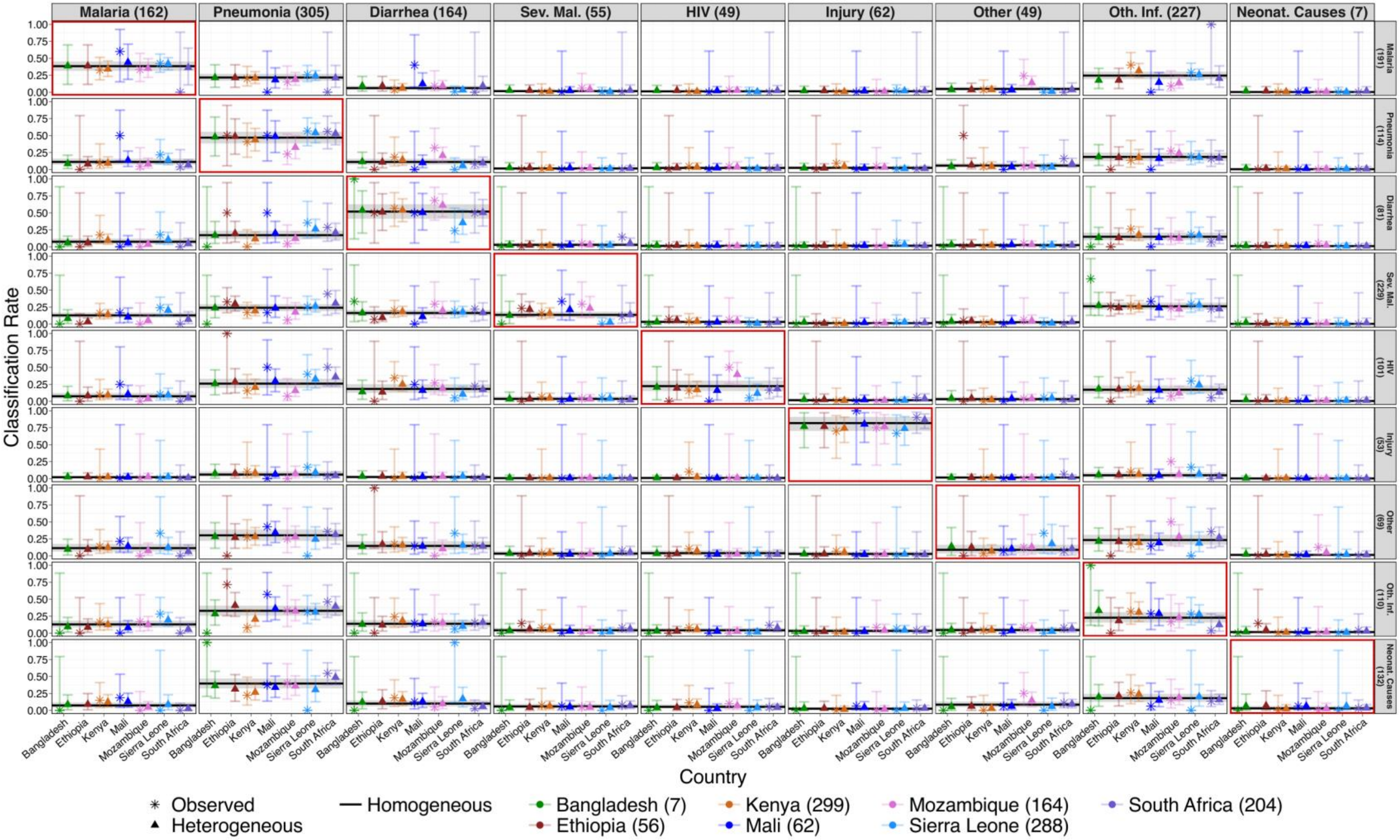


Figure S17. Comparison of observed misclassification rates with in-sample point (posterior mean) and uncertainty (95% credible intervals) estimates from homogeneous and heterogeneous models for InSilicoVA among child (1-59 months) deaths in CHAMPS. Rows and columns correspond to CHAMPS and VA causes, with combined sample sizes across countries indicated in parentheses. Misclassification rates are conditioned on CHAMPS cause (row), so values in each row sum to 1 for each country and method. Sev. mal., Oth. Inf., and Neonat. Causes denote severe malnutrition, other infections, and neonatal causes.

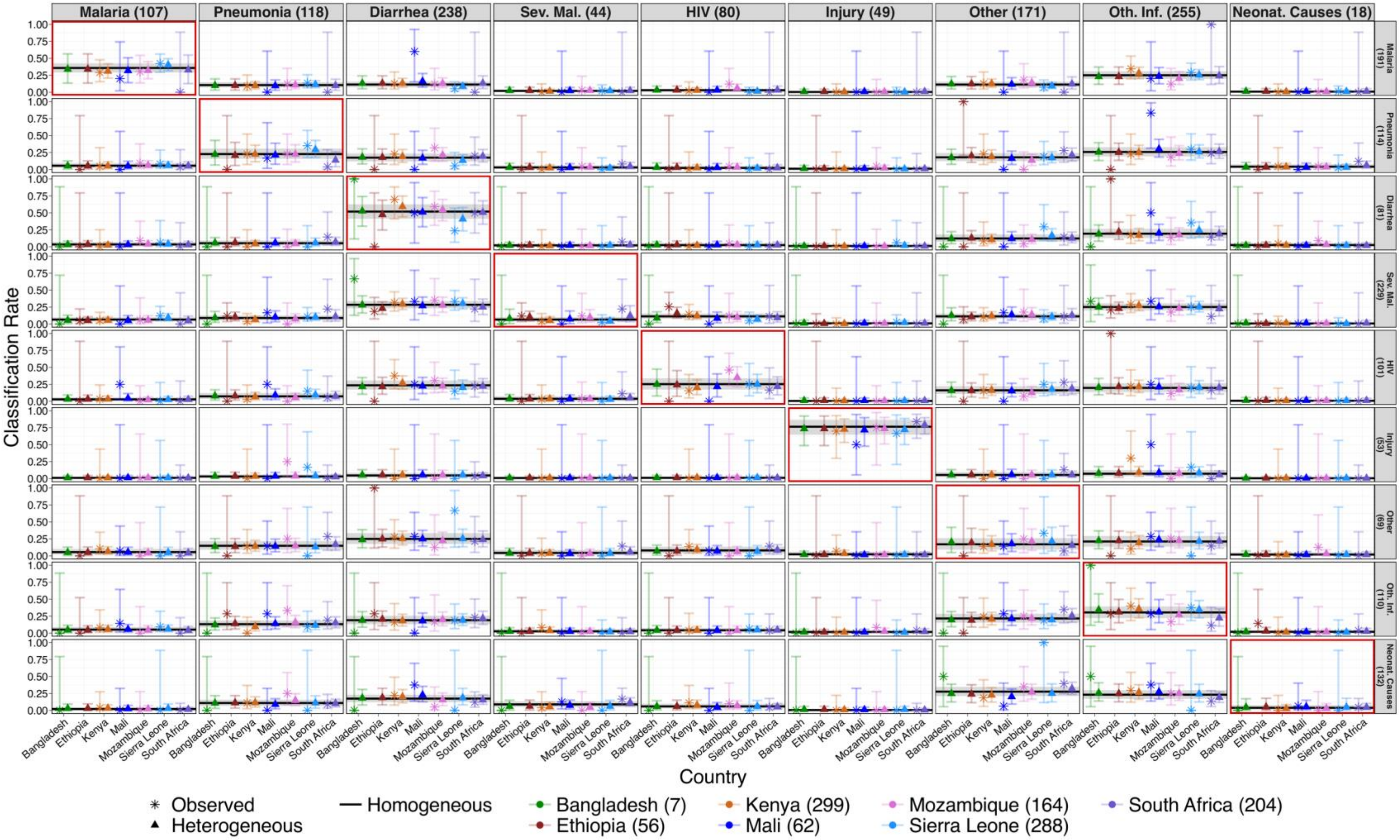


Figure S18. Comparison of observed misclassification rates with in-sample point (posterior mean) and uncertainty (95% credible intervals) estimates from homogeneous and heterogeneous models for InterVA among child (1-59 months) deaths in CHAMPS. Rows and columns correspond to CHAMPS and VA causes, with combined sample sizes across countries indicated in parentheses. Misclassification rates are conditioned on CHAMPS cause (row), so values in each row sum to 1 for each country and method. Sev. mal., Oth. Inf., and Neonat. Causes denote severe malnutrition, other infections, and neonatal causes.

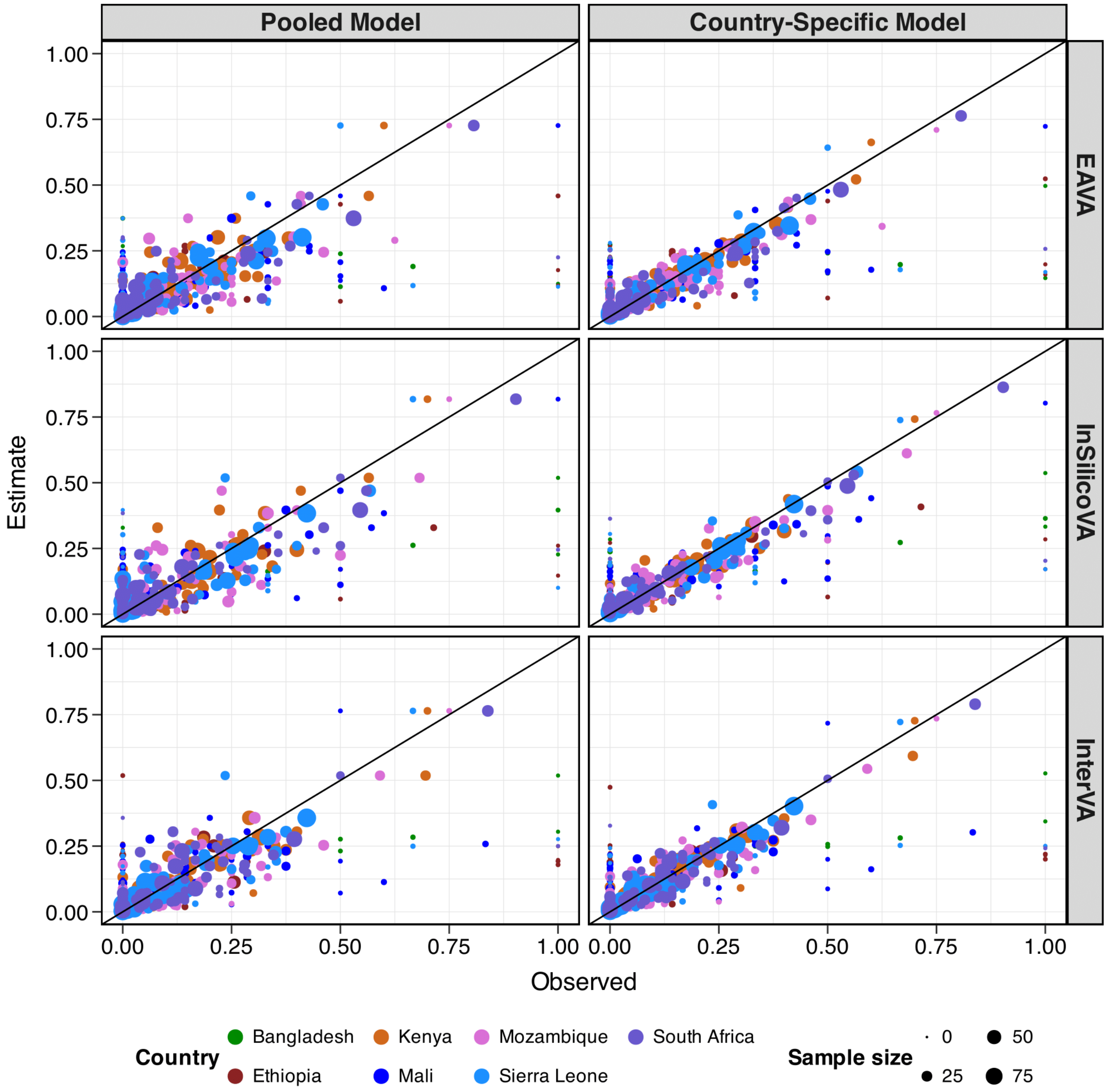


Figure S19. Scatterplots of observed (x-axis) and estimated (y-axis) misclassification rates of EAVA (top row), InSilicoVA (middle row), and InterVA (bottom row) for child (1-59 months) deaths in CHAMPS. It compares estimates from the homogeneous or pooled model (left panels) and country-specific model (right panels). Countries are denoted in different colors. The point sizes reflect the observed sample size for each corresponding CHAMPS cause. The black line corresponds to the $y = x$ line. Compared to the homogeneous model, the country-specific model reduces the average absolute loss with respect to observed rates by 19%, 24%, and 13% for the three algorithms for this age group.

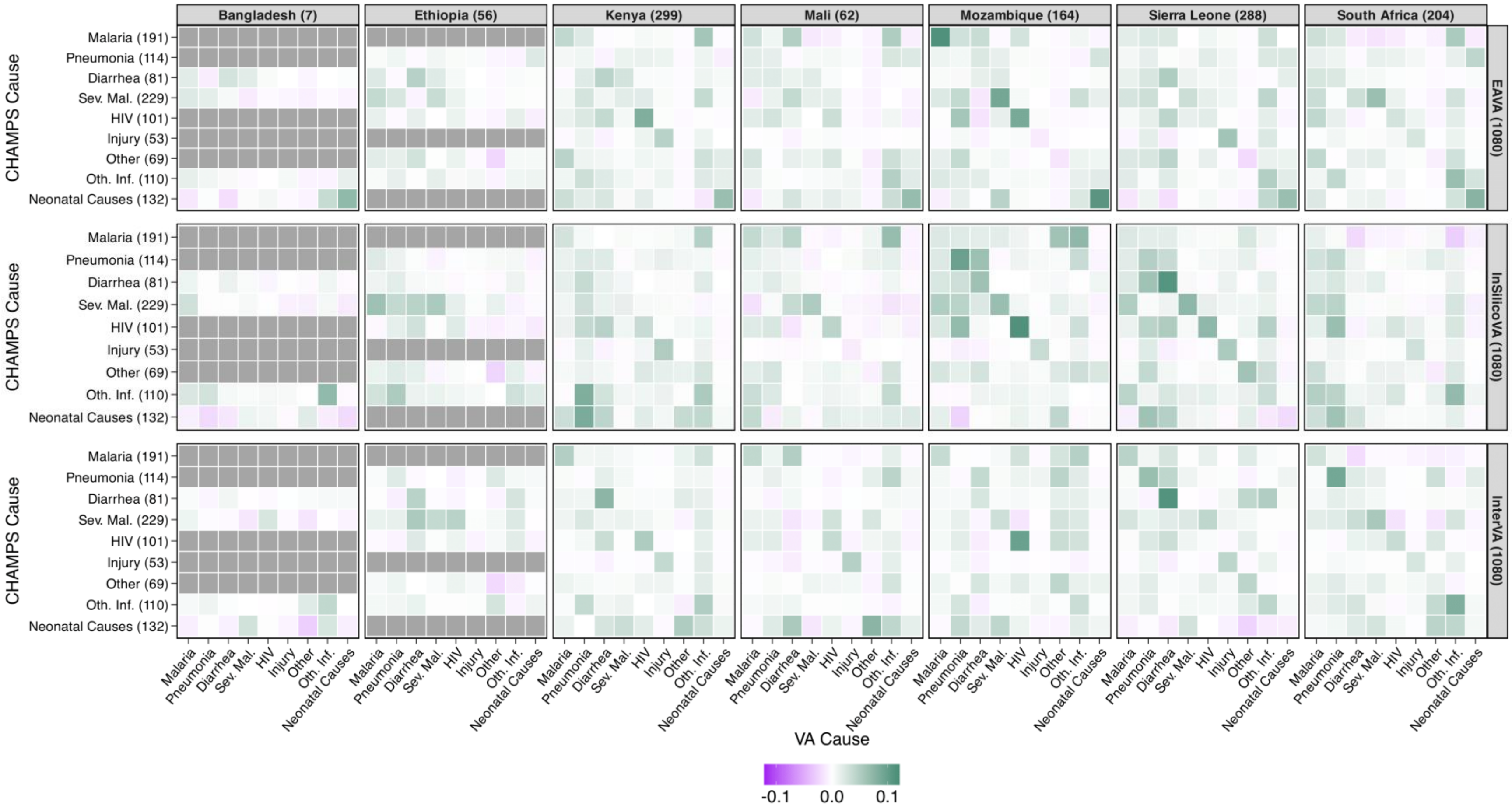


Figure S20. The change in the absolute in-sample bias of point estimates in the country-specific model compared to the homogeneous model among children (1-59 months). Rows and columns are CHAMPS and VA causes. Negative values (green color) are better and indicate a reduction in absolute in-sample bias. Grey cells indicate the CHAMPS cause was not observed for that country. Sev. mal. and Oth. Inf. denote severe malnutrition and other infections. Combined across countries, the country-specific model reduces bias for 69%, 72%, and 69% cause-pairs for EAVA, InSilicoVA, and InterVA with 2-16%, 2-17%, 1-11% improvements (lower bias) for half of them.

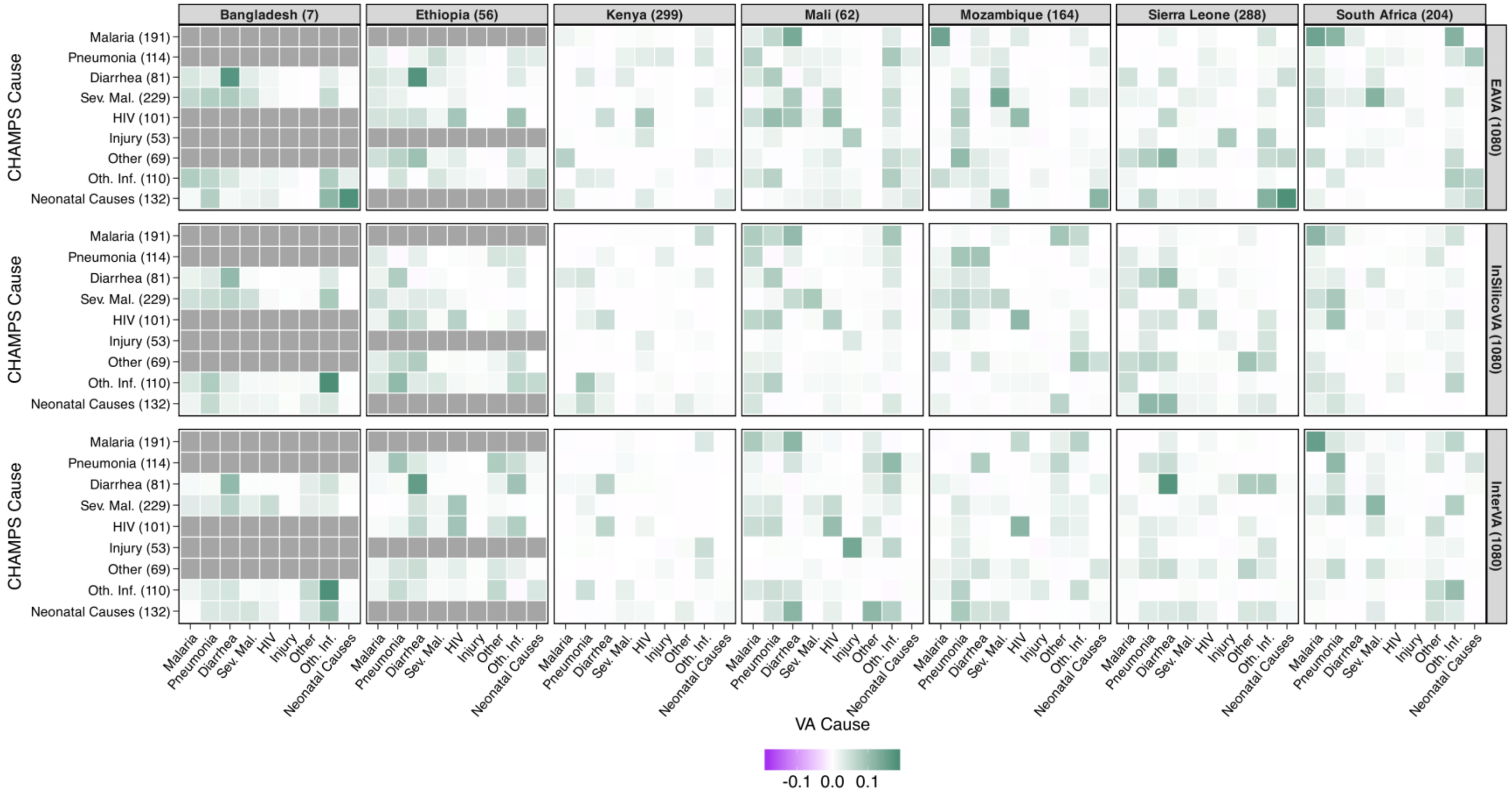


Figure S21. The change in interval scores of 95% credible intervals in the country-specific model compared to the homogeneous model among children (1-59 months). Rows and columns are CHAMPS and VA causes. Negative values (green color) are better and indicate a reduction in absolute interval scores. Grey cells indicate the CHAMPS cause was not observed for that countr. Sev. mal. and Oth. Inf. denote severe malnutrition and other infections. Combined across countries, the country-specific model reduces bias for 65%, 69%, and 69% cause-pairs for EAVA, InSilicoVA, and InterVA with 2-21%, 2-31%, 1-18% improvements (lower scores) for half of them.

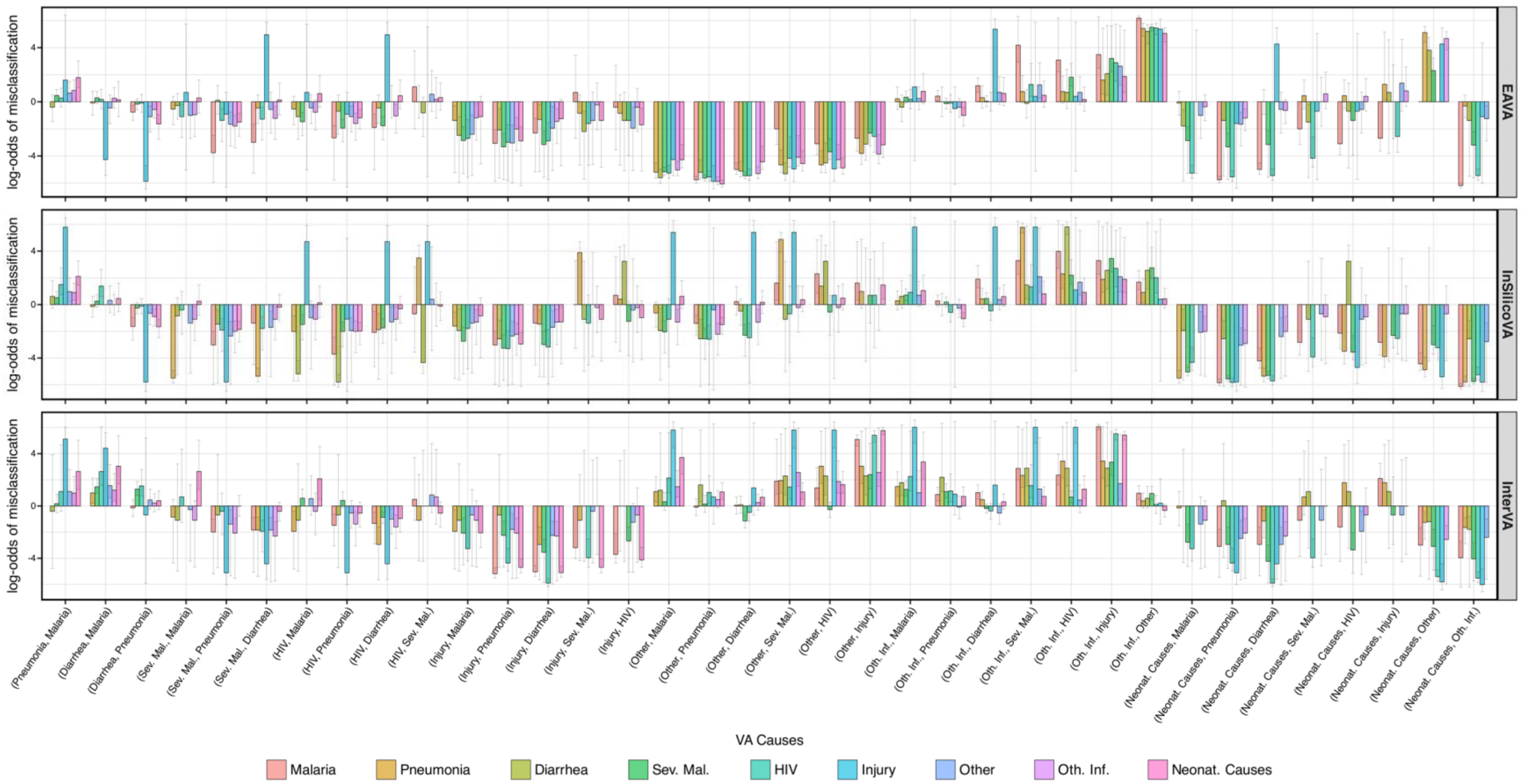


Figure S22. Grouped bar plot showing the similarity of log misclassification odds between CHAMPS causes for child deaths (1-59 months). For each VA cause pair on the horizontal axis, log-odds values for all corresponding CHAMPS causes are represented by color-coded bars on the vertical axis. Error bars indicate 95% confidence intervals calculated from 100,000 bootstrap samples. Sev. mal., Oth. Inf., and Neonat. Causes denote severe malnutrition, other infections, and neonatal causes. The heights of the bars within many groups are similar. This suggests strong evidence favoring the base model proposed in [1]. This is following the log-odds characterization according to Theorem 3.1 from [1].

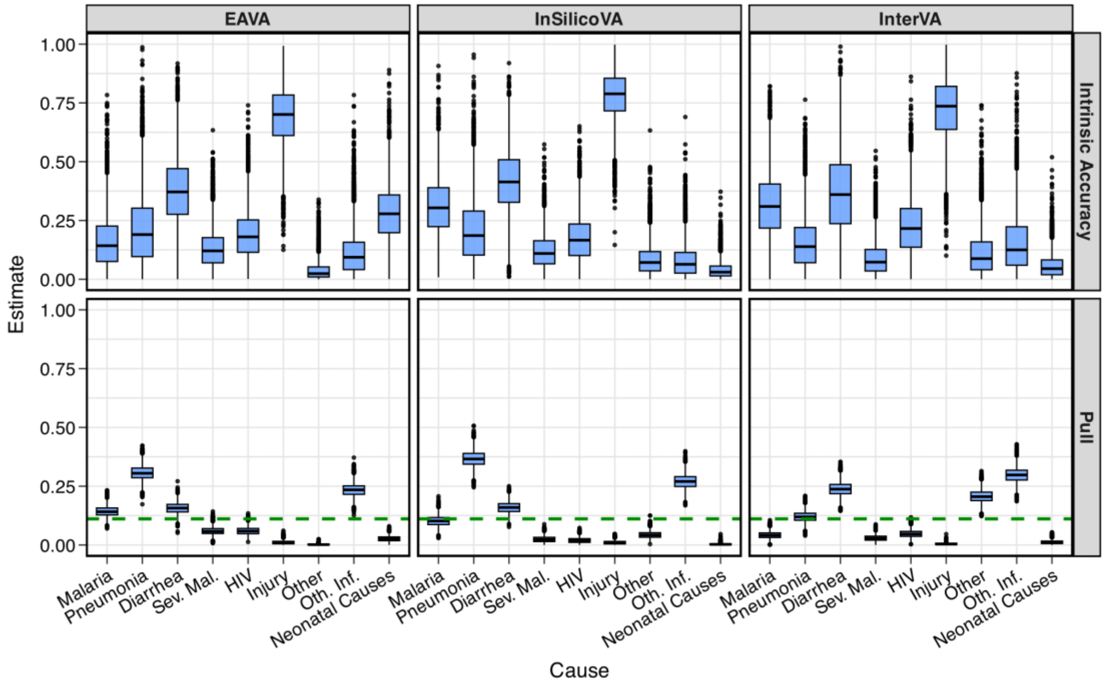


Figure S23. Uncertainty-quantified estimates of intrinsic accuracy and pull for EAVA, InSilicoVA, and InterVA in CHAMPS among child deaths (1-59 months). The horizontal dashed line indicates no preference, corresponding to a uniform pull of 1/9 for each of the nine causes.

**Estimate of intrinsic accuracy and pull.** Figure S23 shows uncertainty-quantified estimates of intrinsic and pull for EAVA, InSilicoVA, and InterVA, the three CCVA algorithms considered here. Accuracy is the highest for injury, with lower accuracies for other and other infections. Without systematic preference, algorithms would misclassify uniformly across causes, resulting in a pull of 1/9 for each of the nine causes (green dashed line). However, the pull estimates indicate systematic biases: EAVA tends to overemphasize pneumonia and other infections while downplaying injury, neonatal causes, and other causes; InSilicoVA similarly overemphasizes pneumonia and other infections while underemphasizing severe malnutrition, HIV, injury, and neonatal causes; InterVA, on the other hand, overstates diarrhea, other infection, and other causes, while minimizing injury, neonatal causes, and severe malnutrition. These highlight the presence of systematic biases in the algorithms.

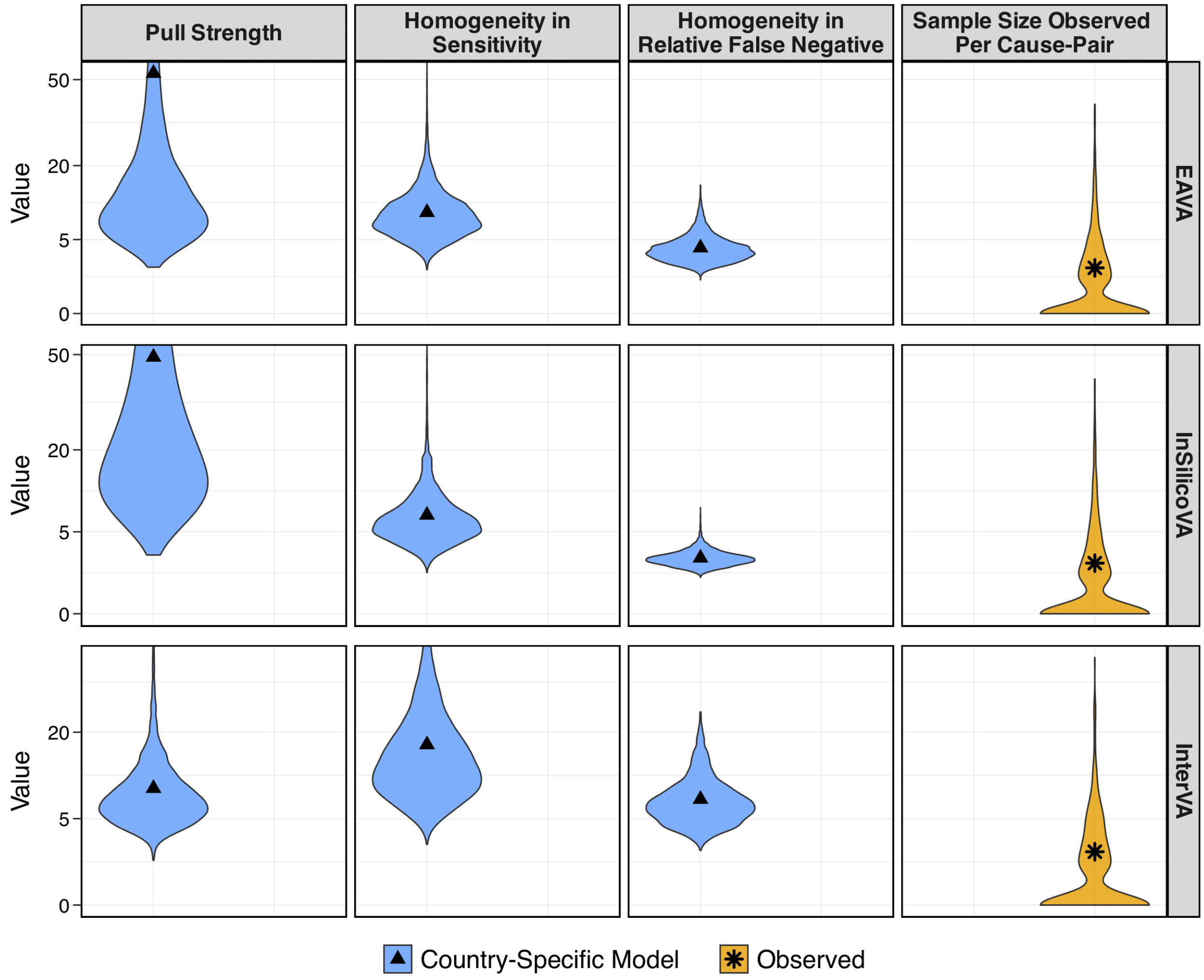


Figure S24. The three panels on the left present uncertainty-quantified estimates of effect sizes in VA misclassification among child (1-59 months) deaths in CHAMPS. The rightmost panel shows the distribution of sample sizes across cause pairs and countries to compare effect size magnitude. Higher values relative to the observed sample size indicate stronger effects. Black points represent the means of each distribution. For clarity, the y-axis is scaled on the square root scale.

Figure S24 presents the estimates of effect sizes (posterior densities; black triangles indicate posterior mean) for pull strength, homogeneity in sensitivity, and homogeneity in relative false negatives from the country-specific model. As outlined in Section S2, we compare effect size estimates (black triangles) with the observed sample sizes (last column; black stars indicate average) across cause pairs and countries for interpretation. First, the posterior mean estimates of pull strength (first column) are 53 for EAVA, 49 for InSilicoVA, and 9 for InterVA, compared to an average observed sample size of 2. This indicates very strong evidence in favor of the base model for EAVA and InSilicoVA, while InterVA reflects the weakest support. Second, the posterior means for the degree of homogeneity in sensitivity (second column) are 9 for EAVA, 7 for InSilicoVA, and 17 for InterVA. Relative to the observed average sample size, this suggests that InterVA exhibits the most homogeneity of sensitivities across countries, with EAVA and InSilicoVA showing moderate heterogeneity. Finally, the estimates for homogeneity in relative false negatives (third column) are 4 for EAVA, 2 for InSilicoVA, and 7 for InterVA. These low values, compared to the observed sample sizes, indicate a higher level of heterogeneity in relative false negatives than in sensitivity—particularly pronounced in InSilicoVA and InterVA, and somewhat less so in EAVA. As for neonates, overall these results highlight differing levels of systematic structure and heterogeneity in the misclassification patterns of each CCVA algorithm and demonstrate the misclassification model’s adaptive capacity to account for these differences, enabling more accurate estimation of the misclassification matrices.

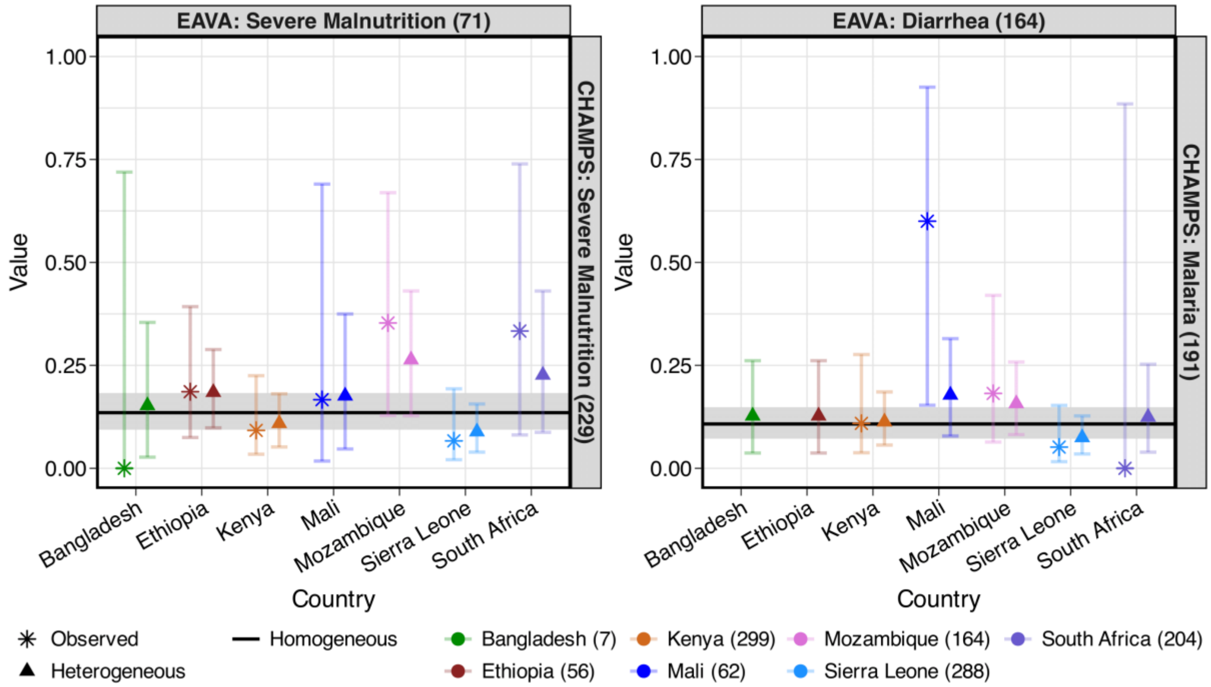


Figure S25. Left panel: Observed and estimated sensitivities of EAVA for severe malnutrition across CHAMPS countries for child deaths (1-59 months). Right panel: Observed and estimated false negative rates of EAVA for CHAMPS cause malaria and VA cause diarrhea across CHAMPS countries for child deaths.

**Examining the Causes of Heterogeneity in VA Misclassification.** The left panel of Figure S25 shows the sensitivity of severe malnutrition by EAVA. There are four sets of severe malnutrition diagnostic criteria in the EAVA hierarchy for 1-59 months old children, depending on whether the child also had diarrhea/dysentery, pneumonia, possible diarrhea/dysentery/pneumonia, or only malnutrition - which is placed at the bottom of the hierarchy. Kenya and Sierra Leone show lower sensitivity compared to other CHAMPS countries. In Kenya, 40 out of the 76 records that have an underlying cause of severe malnutrition meet one of the sets of diagnostic criteria for severe malnutrition by EAVA. However, only seven records have severe malnutrition as their top cause in the hierarchy. In Sierra Leone, 32 out of 75 of the records that are severe malnutrition by CHAMPS cause meet the diagnostic criteria for one of the forms of severe malnutrition diagnosed by EAVA. However, only five records have severe malnutrition as their top cause in the EAVA hierarchy. Like what was observed in neonates, a multi-cause analysis may improve the sensitivity of some causes that are lower in the EAVA hierarchy in 1-59-month-old children.

#### S3.2 Case Study: CSMF in Mozambique Using COMSA-Mz Data

**Raw CSMF estimates.** The blue bars in Figure S27 represent uncalibrated CSMF estimates for various causes based on VA-only COD predictions from EAVA, InSilicoVA, and InterVA in COMSA-Mz. The estimates differ considerably across the algorithms. Among the 2812 child deaths in COMSA, EAVA identifies other infections as the most common cause of death, while InSilicoVA and InterVA identify diarrhea as the leading cause. For the second most common cause, EAVA predicts pneumonia, whereas InSilicoVA and InterVA indicate other infections.

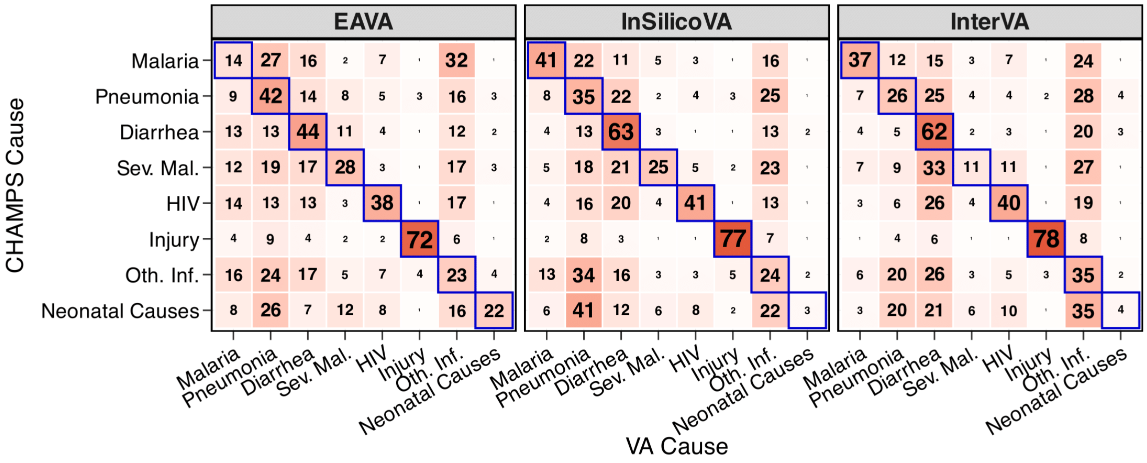


Figure S26. Obtained from CHAMPS analysis of child (1-59 months) deaths, this is the (expected) misclassification (without 'other') for Mozambique that is used as an informative prior in the modular VA-calibration. Sensitivities are along diagonals (outlined in blue). Sev. mal. and Oth. Inf. denote severe malnutrition and other infections.

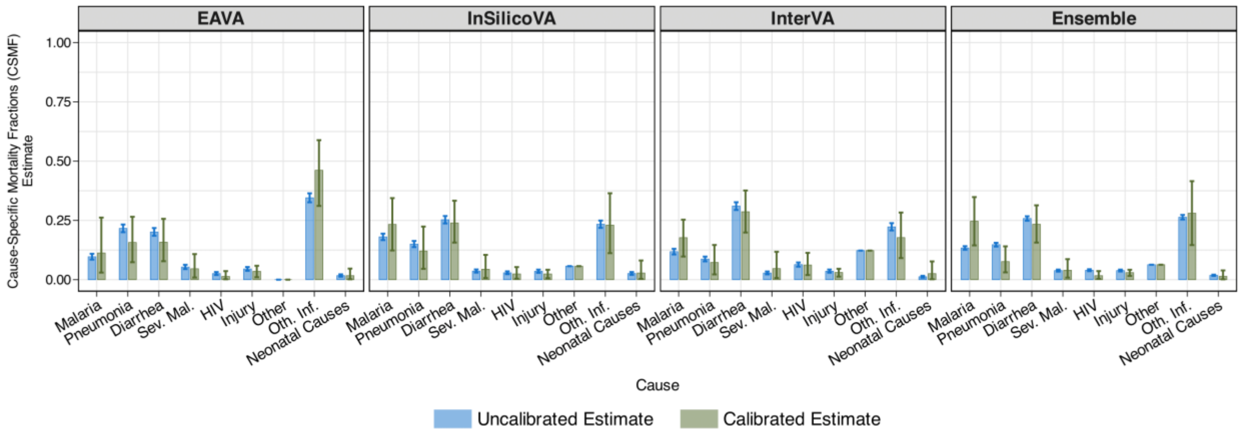


Figure S27. Comparison of uncalibrated (blue) and calibrated (green) Cause-Specific Mortality Fraction (CSMF) estimates for children (1-59 months) using EAVA, InSilicoVA, InterVA, and their ensemble. Bar heights represent the point estimates (posterior means), while the error bars indicate uncertainty (95% credible intervals). Although uncalibrated CSMF estimates show lower uncertainty than calibrated ones, they assume perfect classification—an assumption contradicted by substantial VA misclassification observed in CHAMPS. No calibration can lead to overconfidence and biased CSMF estimates. Sev. mal. and Oth. Inf. denote severe malnutrition and other infections.

**Calibrated CSMF estimates.** We calibrate the raw CSMF estimates based on COMSA-Mz data, using the Mozambique-specific estimates of misclassification rates for children (1-59 months) for each CCVA algorithm as presented in Figure S26. This adjusts for misclassification and produces uncertainty-quantified, calibrated CSMF estimates (in green) in Figure S27.

The calibration effects on CSMFs somewhat vary across the algorithms. For EAVA, the CSMFs for pneumonia and diarrhea decrease, while the CSMF for other infections increases. We find that 14% and 16% of child deaths from pneumonia are misclassified as diarrhea and other infections, respectively; 13% of deaths from diarrhea are misclassified as pneumonia; and 24% and 17% of deaths from other infections are misclassified as pneumonia and diarrhea (see Figure S26). Given that pneumonia, diarrhea, and other infections account for 76% of child deaths in COMSA-Mz and other infections have the lowest sensitivity among the three, VA calibration results in an increased CSMF for other infections and decreased CSMFs for pneumonia and diarrhea. For InSilicoVA, the CSMF for pneumonia decreases, while the CSMF for malaria increases. Finally, in contrast to EAVA, the calibrated CSMF for InterVA shows a decrease in other infections but an increase in malaria. We see from Figure S26 that, for InterVA, 24% of malaria deaths are misclassified as other infections, while only 6% of deaths from other infections are misclassified as malaria. This increases the CSMF of malaria and decreases the CSMF of other infections for InterVA after calibration.

The final calibrated ensemble CSMF estimates, shown in the far-right panel of Figure S27, attribute 28% of child deaths to other infections, 25% to malaria, 24% to diarrhea, 7% to pneumonia, 6% to other causes, 4% to severe malnutrition, 3% to injury, 2% to HIV, and 1% to neonatal causes. The 95% Bayesian credible intervals for these calibrated CSMFs help identify causes that significantly differ from the uncalibrated estimates. For the ensemble, the calibrated CSMF interval for malaria (15-35%) is higher than its uncalibrated estimate (13%), indicating an increase post-calibration. Conversely, for pneumonia, the interval (314%) is lower than its uncalibrated estimate (15%), indicating a decrease. For other causes, the credible intervals either overlap with or are close to the uncalibrated estimates. The results for malaria and pneumonia align with findings by [2], while the current analysis indicates no substantial change for other infections after calibration.

#### S4. CSMF Estimates in Mozambique Using COMSA-Mozambique

Table S1. Uncalibrated and calibrated estimates (in percentage) of cause-specific mortality fractions (CSMF) for neonates (0-27 days) in Mozambique, derived based on the EAVA, InSilicoVA, InterVA algorithms and their ensemble, using data from the COMSA-Mozambique study. Each cell displays the point estimate (posterior mean), with its uncertainty (95% credible interval) provided in parentheses. Con. mal., Sep./Menin./Inf., IPRE denote congenital malformation, sepsis/meningitis/infection, and intrapartum-related events. Calibration is not performed for the 'other' cause.

| Algorithm | Calibration Type | Con. Mal. | Pneumonia | Sep./Menin./Inf. | IPRE | Other | Prematurity |
|---|---|---|---|---|---|---|---|
| EAVA | Uncalibrated | 5 (4, 7) | 19 (16, 22) | 29 (26, 32) | 24 (22, 27) | 4 (4, 4) | 19 (16, 21) |
| | Calibrated | 4 (1, 10) | 24 (7, 45) | 37 (16, 56) | 19 (8, 34) | 4 (4, 4) | 13 (5, 23) |
| InSilicoVA | Uncalibrated | 0 (0, 1) | 12 (10, 14) | 31 (29, 34) | 27 (25, 30) | 5 (5, 5) | 24 (22, 27) |
| | Calibrated | 3 (0, 11) | 10 (2, 25) | 50 (37, 63) | 21 (10, 31) | 5 (5, 5) | 11 (5, 19) |
| InterVA | Uncalibrated | 4 (3, 5) | 7 (5, 8) | 17 (15, 20) | 24 (21, 26) | 7 (7, 7) | 42 (39, 45) |
| | Calibrated | 3 (0, 8) | 8 (1, 21) | 26 (9, 45) | 12 (5, 22) | 7 (7, 7) | 44 (26, 62) |
| Ensemble | Uncalibrated | 3 (2, 3) | 12 (11, 13) | 26 (24, 27) | 25 (24, 27) | 5 (5, 5) | 29 (28, 31) |
| | Calibrated | 2 (0, 6) | 9 (2, 21) | 57 (46, 67) | 14 (6, 22) | 5 (5, 5) | 12 (6, 19) |

Table S2. Uncalibrated and calibrated estimates (in percentage) of cause-specific mortality fractions (CSMF) for children (1-59 months) in Mozambique, derived based on the EAVA, InSilicoVA, InterVA algorithms and their ensemble, using data from the COMSA-Mozambique study. Each cell displays the point estimate (posterior mean), with its uncertainty (95% credible interval) provided in parentheses. Sev. mal. and Oth. Inf. denote severe malnutrition and other infections. Calibration is not performed for the 'other' cause.

| Algorithm | Calibration Type | Malaria | Pneumonia | Diarrhea | Sev. Mal. | HIV | Injury | Other | Oth. Inf. | Neonatal Causes |
|---|---|---|---|---|---|---|---|---|---|---|
| EAVA | Uncalibrated | 10 (8, 11) | 22 (20, 23) | 20 (19, 22) | 5 (4, 6) | 3 (2, 3) | 5 (4, 5) | 0 (0, 0) | 35 (33, 36) | 2 (1, 2) |
| | Calibrated | 11 (3, 26) | 16 (7, 27) | 16 (8, 26) | 5 (1, 11) | 1 (0, 4) | 4 (1, 6) | 0 (0, 0) | 46 (31, 59) | 2 (0, 5) |
| InSilicoVA | Uncalibrated | 18 (17, 19) | 15 (14, 16) | 25 (24, 27) | 4 (3, 4) | 3 (2, 4) | 4 (3, 4) | 6 (6, 6) | 23 (22, 25) | 3 (2, 3) |
| | Calibrated | 23 (12, 34) | 12 (5, 22) | 24 (16, 33) | 4 (1, 10) | 2 (0, 5) | 2 (1, 4) | 6 (6, 6) | 23 (11, 36) | 3 (0, 8) |
| InterVA | Uncalibrated | 12 (11, 13) | 9 (8, 10) | 31 (29, 33) | 3 (2, 3) | 6 (5, 7) | 4 (3, 4) | 12 (12, 12) | 22 (21, 24) | 1 (1, 2) |
| | Calibrated | 18 (10, 25) | 7 (2, 15) | 29 (20, 38) | 5 (1, 12) | 6 (2, 11) | 3 (1, 5) | 12 (12, 12) | 18 (9, 28) | 3 (0, 8) |
| Ensemble | Uncalibrated | 13 (13, 14) | 15 (14, 16) | 26 (25, 27) | 4 (3, 4) | 4 (4, 4) | 4 (3, 4) | 6 (6, 6) | 26 (25, 27) | 2 (2, 2) |
| | Calibrated | 25 (14, 35) | 8 (3, 14) | 23 (16, 31) | 4 (1, 9) | 2 (0, 4) | 3 (2, 4) | 6 (6, 6) | 28 (15, 42) | 1 (0, 4) |
